# Decoding the diet–gut–liver axis: links between dietary pattern adherence, gut microbiome, and hepatic health

**DOI:** 10.64898/2026.05.04.26352208

**Authors:** Keyong Deng, Quinten R. Ducarmon, Anastasia Godneva, Zheqing Zhang, Astrid van Hylckama Vlieg, Frits R. Rosendaal, Georg Zeller, Eran Segal, Ruifang Li-Gao, DIYUFOOD consortium

## Abstract

Metabolic dysfunction-associated steatotic liver disease (MASLD) is rapidly becoming the leading cause of chronic liver disease and confers substantial cardiometabolic burden. Diet quality and gut microbiota composition have been implicated in MASLD development; however, the interplay among diet, gut microbiota, and hepatic health remains insufficiently characterized. Here, in 9,616 deeply phenotyped middle-aged participants (mean age 52 years) from the Human Phenotype Project, we investigated how five dietary quality indices capturing complementary dimensions of healthy eating, including plant-based (hPDI), Mediterranean-style (AMED), anti-inflammatory (rDII), anti-hyperinsulinemic (rEDIH), and overall quality (AHEI), relate to gut microbial composition and liver steatosis. Dietary pattern scores were derived from two-week continuous diet logs, gut microbiota was characterized by shotgun metagenomic sequencing, and hepatic health was assessed by both ultrasound-derived metrics and prevalent MASLD status. Adherence to each of the five healthy dietary patterns was inversely associated with MASLD prevalence and positively associated with liver speed of sound (SoS), an ultrasound-derived metric that correlates inversely with hepatic fat content. Across all five dietary patterns, greater adherence was consistently associated with 138 gut microbial species, including inverse associations with *Flavonifractor plautii*, *Dysosmobacter welbionis*, *Ruthenibacterium lactatiformans*, *Bilophila wadsworthia*, and *Phocea massiliensis*. These five species were also associated with lower liver SoS and higher odds of prevalent MASLD, emerging as potential mediators of the diet–liver relationship in cross-sectional mediation analyses after adjustment for body mass index (BMI). This study identifies candidate microbial targets for future interventional studies investigating dietary strategies for MASLD prevention.

## INTRODUCTION

Metabolic dysfunction-associated steatotic liver disease (MASLD) is one of the major global health burdens, with a rising prevalence estimated at 25–30% worldwide^1^. Its clinical spectrum ranges from simple steatosis to steatohepatitis, fibrosis, cirrhosis, and hepatocellular carcinoma, and MASLD has been implicated in the development of extrahepatic disorders, including type 2 diabetes (T2D) and cardiovascular disease (CVD)^1–3^. Given that MASLD typically progresses without overt symptoms, population-level strategies targeting modifiable risk factors for early prevention remain a critical unmet need.

Converging evidence indicates that improved diet quality is a cornerstone of MASLD management^4–6^. Dietary pattern scores, which capture overall dietary habits, provide a comprehensive and clinically relevant diet assessment^7,8^. Adherence to healthy dietary patterns, as quantified by the Alternate Mediterranean Diet score (AMED), Alternative Healthy Eating Index (AHEI), or healthy Plant-based Diet Index (hPDI), has separately been associated with reduced liver fat accumulation and MASLD risk^4,5,9–13^. Conversely, habitual diets with inflammatory or hyperinsulinemic potential, quantified by the Dietary Inflammatory Index (DII) and Empirical Dietary Index for Hyperinsulinemia (EDIH), have been associated with increased MASLD risk^14–17^. Yet, few studies have systematically compared these diet–liver associations within a single analytical framework, an approach that could identify which dietary patterns are most relevant to hepatic health. Furthermore, the growing availability of non-invasive tools such as two-dimensional shear wave elastography (2D-SWE) raises the question of whether healthy diet adherence is associated not only with binary MASLD status but also with continuous measures of hepatic tissue properties.

The pathogenesis of MASLD is known to be multifaceted, involving complex interactions among genetic, environmental, and metabolic risk factors^1,2^. The gut microbiome is increasingly recognized as a contributor to MASLD development through modulation of biological processes such as cholesterol metabolism, bile acid signaling, and immune activation^18–21^. Disruption of gut microbial communities has been linked to increased intestinal permeability and translocation of microbial products (e.g., lipopolysaccharides) to the liver, thereby promoting hepatic inflammation^22–25^. Diet is a major determinant of gut microbial composition and function^26–28^, shaping the production of metabolites (e.g., short-chain fatty acids and secondary bile acids) that reach the liver via the portal circulation and influence host metabolic health. Growing evidence indicates that gut microbial species associated with healthy dietary habits are also linked to favorable metabolic profiles and health outcomes^29–32^. However, it remains unclear whether adherence to different healthy dietary patterns is associated with shared or distinct gut microbial signatures and, if so, whether these signatures are related to hepatic health.

In the current study, we leveraged the deeply phenotyped Human Phenotype Project (HPP)^33^, which integrates dietary logs, gut metagenomics, self-reported MASLD status, and non-invasive hepatic imaging by ultrasound and 2D-SWE that captures fibrosis-, inflammation-, and steatosis-related tissue properties. We evaluated five dietary pattern scores (AHEI, hPDI, AMED, reversed DII [rDII], and reversed EDIH [rEDIH]) based on 2-week continuous diet logs, harmonized such that higher values denote greater adherence to healthy dietary patterns. We aimed to (1) examine the associations between different dietary pattern scores and prevalent MASLD and ultrasound-derived hepatic metrics; (2) identify gut microbial signatures associated with healthy diet adherence; and (3) explore whether the abundance of gut microbial species may partially account for the relationship between healthy diet adherence and hepatic health (**Figure 1**).

**Figure 1.**
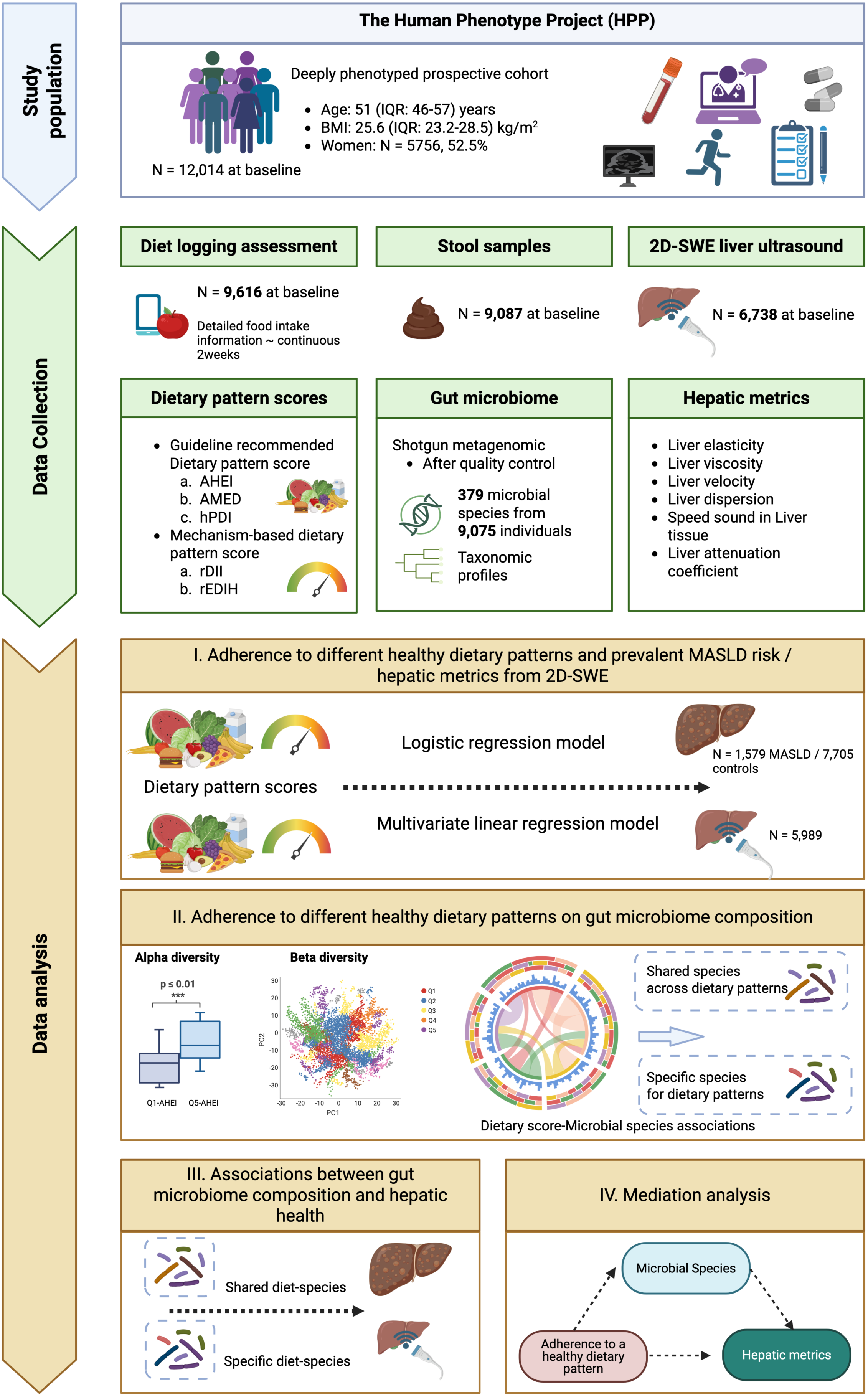
Schematic of the study workflow. At baseline, a total of 12,014 individuals enrolled in the Human Phenotype Project (HPP). 9,616 individuals had valid dietary logging data. A total of 9,087 individuals had valid gut metagenomic measures. A total of 6,738 individuals underwent liver ultrasound and two-dimensional shear wave elastography examination. We computed five different energy-adjusted dietary pattern scores (three guideline-recommended indices: AHEI, AMED, hPDI and two mechanism-based indices: rDII and rEDIH) to indicate individuals’ healthy diet adherence. Six hepatic metrics (liver elasticity, viscosity, velocity, dispersion, attenuation coefficient, and liver speed of sound) were included. Logistic regression models and generalized linear models (GLMs) were employed to investigate the associations of healthy diet adherence with prevalent MASLD and hepatic metrics, respectively. In addition, a microbiome-wide association analyses for adherence to different healthy dietary patterns was conducted at species level. Microbial species associated with healthy diet adherence were further examined for associations with prevalent MASLD or hepatic metrics. Lastly, a mediation analysis was conducted to identify microbial species as candidate mediators that potentially account for the diet–liver association. AHEI, Alternative Healthy Eating Index; AMED, Alternate Mediterranean Diet Score; BMI, body mass index; CVD, cardiovascular disease; FDR, false discovery rate; hPDI, healthy Plant-based Diet Index; MASLD, Metabolic dysfunction-associated steatotic liver disease; rDII, reversed Dietary Inflammatory Index; rEDIH, reversed Empirical Dietary Index for Hyperinsulinemia. The figure was created using *BioRender*.

## RESULTS

### Characteristics of the study population

**Table 1** presents the baseline characteristics of 9,616 individuals with complete and valid dietary log data for a median of 14 (interquartile range [IQR]: 13–14) consecutive days. Participants had a mean age of 51.9 (standard deviation [SD]: 7.8) years and an average body mass index (BMI) of 26.1 (SD: 4.1) kg/m^2^, and 5,044 (52.5%) were women. The five energy-adjusted dietary pattern scores (AHEI, AMED, hPDI, rDII, and rEDIH) were approximately normally distributed and positively correlated with each other (Spearman’s ρ = 0.17 to 0.76), although rEDIH correlated less strongly with the other four scores (ρ < 0.5) (**Supplementary Figures 1 and 2**). Across all dietary pattern scores, individuals in the highest quintile were older, more educated, more physically active, slightly leaner, and more likely to use vitamins, but were less likely to smoke or use nonsteroidal anti-inflammatory drugs (NSAIDs) or aspirin, than those in the lowest quintile. Women predominated in the highest quintile of AHEI, AMED, hPDI, and rEDIH, whereas men predominated in the highest quintile of rDII (**Table 1**).

**Table 1.** Baseline characteristics in the lowest and highest quintiles of energy-adjusted dietary pattern scores of enrolled participants.

| Dietary pattern scores | Overall | AHEI |  | AMED |  | hPDI |  | rDII |  | rEDIH |  |
| --- | --- | --- | --- | --- | --- | --- | --- | --- | --- | --- | --- |
|  | N = 9616 | Q1<br>N = 1924 | Q5<br>N = 1923 | Q1<br>N = 1924 | Q5<br>N = 1923 | Q1<br>N = 1924 | Q5<br>N = 1923 | Q1<br>N = 1924 | Q5<br>N = 1923 | Q1<br>N = 1924 | Q5<br>N = 1923 |
| Logging days, median (IQR) | 14 (13,14) | – | – | – | – | – | – | – | – | – | – |
| Energy-adjusted scores, mean (SD) | – | 36.5 (5.0) | 65.6 (4.1) | 1.6 (0.7) | 6.6 (0.6) | 42.3 (2.5) | 60.0 (3.3) | -2.4 (0.3) | 0.2 (0.5) | -0.7 (0.1) | -0.1 (0.1) |
| Female, n (%) | 5044 (52.5) | 662 (34.4) | 1218 (63.3) | 901 (46.8) | 1168 (60.7) | 792 (41.2) | 1179 (61.3) | 1107 (57.5) | 841 (43.7) | 491 (25.5) | 1359 (70.7) |
| Age at baseline, years, mean (SD) | 51.9 (7.8) | 49.8 (7.3) | 53.6 (7.8) | 50.1 (7.4) | 53.7 (7.9) | 50.0 (7.4) | 53.4 (7.9) | 49.5 (7.1) | 53.4 (8.0) | 49.8 (7.3) | 52.7 (7.9) |
| Missing, n (%) | 637 (6.6) | 148 (7.7) | 99 (5.1) | 152 (7.9) | 94 (4.9) | 125 (6.5) | 133 (6.9) | 134 (7.0) | 131 (6.3) | 141 (7.3) | 110 (5.7) |
| BMI, kg/m², mean (SD) | 26.1 (4.1) | 27.0 (4.2) | 25.3 (4.0) | 26.6 (4.3) | 25.4 (4.0) | 26.6 (4.2) | 25.1 (3.9) | 26.5 (4.4) | 25.6 (4.1) | 27.1 (4.1) | 25.0 (4.1) |
| Missing | 659 (6.9) | 154 (8.0) | 106 (5.5) | 156 (8.1) | 100 (5.2) | 132 (6.9) | 135 (7.0) | 141 (7.3) | 125 (6.5) | 146 (7.6) | 115 (6.0) |
| Smoking status, n (%) |  |  |  |  |  |  |  |  |  |  |  |
| Current | 1012 (10.5) | 240 (12.5) | 152 (7.9) | 249 (12.9) | 157 (8.2) | 223 (11.6) | 173 (9.0) | 272 (14.1) | 145 (7.5) | 202 (10.5) | 191 (9.9) |
| Former | 862 (9.0) | 170 (8.8) | 173 (9.0) | 176 (9.1) | 183 (9.5) | 162 (8.4) | 173 (9.0) | 169 (8.8) | 178 (9.3) | 200 (10.4) | 156 (8.1) |
| Never | 5543 (57.6) | 1039 (54.0) | 1175 (61.1) | 1025 (53.3) | 1178 (61.3) | 1084 (56.3) | 1163 (60.5) | 1031 (53.6) | 1201 (62.5) | 1078 (56.0) | 1161 (60.4) |
| Missing | 2199 (22.9) | 475 (24.7) | 423 (22.0) | 474 (24.6) | 405 (21.1) | 455 (23.6) | 414 (21.5) | 452 (23.5) | 399 (20.7) | 444 (23.1) | 415 (21.6) |
| Physical activity, MET-h/day, median (IQR) | 11.6 (4.4,23.1) | 9.9 (3.3,21.1) | 12.9 (5.7,25.0) | 10.2 (3.3,21.6) | 12.3 (5.3,24.5) | 10.2 (3.3,21.2) | 13.1 (5.5,25.1) | 10.2 (3.4,21.2) | 12.3 (5.0,24.3) | 9.9 (3.6,20.5) | 12.9 (5.2,24.9) |
| Missing, n (%) | 2218 (22.0) | 461 (24.0) | 399 (20.7) | 457 (23.8) | 384 (20.0) | 435 (22.6) | 390 (20.3) | 432 (22.5) | 381 (19.8) | 427 (22.2) | 396 (20.6) |
| Education status, n (%) |  |  |  |  |  |  |  |  |  |  |  |
| High | 6715 (69.8) | 1310 (68.1) | 1360 (70.7) | 1306 (67.9) | 1372 (71.3) | 1333 (69.3) | 1350 (70.2) | 1333 (69.3) | 1374 (71.5) | 1357 (70.5) | 1350 (70.2) |
| Low | 692 (7.2) | 137 (7.1) | 143 (7.4) | 146 (7.6) | 142 (7.4) | 141 (7.3) | 162 (8.4) | 139 (7.2) | 148 (7.7) | 128 (6.7) | 159 (8.3) |
| Missing | 2209 (23.0) | 477 (24.8) | 420 (21.8) | 472 (24.5) | 409 (21.3) | 450 (23.4) | 411 (21.4) | 452 (23.5) | 401 (20.9) | 439 (22.8) | 414 (21.5) |
| Alcohol intake, g/day, median (IQR) | 0.6 (0.0,2.9) | 0.5 (0.0,2.2) | 0.8 (0.0,4.2) | 0.6 (0.0,2.6) | 0.6 (0.0,3.0) | 0.6 (0.0,3.0) | 0.5 (0.0,2.5) | 0.8 (0.0,3.2) | 0.5 (0.0,2.5) | 0.8 (0.0,3.2) | 0.5 (0.0,2.7) |
| Energy intake, kcal/day, mean (SD) | 1605.8 (319.2) | 1671.1 (314.0) | 1588.8 (311.2) | 1604.1 (329.4) | 1589.5 (288.7) | 1658.3 (318.9) | 1607.9 (323.3) | 1671.2 (293.8) | 1683.6 (313.9) | 1698.9 (311.0) | 1605.5 (328.3) |
| Sleep hours, mean (SD) | 6.7 (0.9) | 6.7 (1.0) | 6.8 (0.9) | 6.7 (1.0) | 6.8 (0.9) | 6.7 (0.9) | 6.8 (0.9) | 6.8 (1.0) | 6.7 (0.9) | 6.7 (0.9) | 6.8 (0.9) |
| Missing, n (%) | 2234 (23.2) | 479 (24.9) | 430 (22.4) | 480 (24.9) | 411 (21.4) | 461 (24.0) | 421 (21.9) | 458 (23.8) | 412 (21.4) | 448 (23.3) | 420 (21.8) |
| Vitamin use, n (%) |  |  |  |  |  |  |  |  |  |  |  |
| Use | 1788 (18.6) | 214 (11.1) | 485 (25.2) | 237 (12.3) | 458 (23.8) | 241 (12.5) | 513 (26.7) | 299 (15.5) | 448 (23.3) | 244 (12.7) | 485 (25.2) |
| No use | 7228 (75.2) | 1569 (81.5) | 1336 (69.5) | 1549 (80.5) | 1365 (71.0) | 1569 (81.5) | 1302 (67.7) | 1490 (77.4) | 1349 (70.2) | 1535 (79.8) | 1332 (69.3) |
| Missing | 600 (6.2) | 141 (7.3) | 102 (5.3) | 138 (7.2) | 100 (5.2) | 114 (5.9) | 108 (5.6) | 135 (7.0) | 126 (6.6) | 145 (7.5) | 106 (5.5) |
| Hormone use, n (%) |  |  |  |  |  |  |  |  |  |  |  |
| Use | 981 (10.2) | 125 (6.5) | 252 (13.1) | 158 (8.2) | 235 (12.2) | 155 (8.1) | 245 (12.7) | 205 (10.7) | 167 (8.7) | 102 (5.3) | 244 (12.7) |
| No use | 8035 (83.6) | 1658 (86.2) | 1569 (81.6) | 1628 (84.6) | 1588 (82.6) | 1655 (86.0) | 1570 (81.6) | 1584 (82.3) | 1630 (84.8) | 1677 (87.2) | 1573 (81.8) |
| Missing | 600 (6.2) | 141 (7.3) | 102 (5.3) | 138 (7.2) | 100 (5.2) | 114 (5.9) | 108 (5.6) | 135 (7.0) | 126 (6.6) | 145 (7.5) | 106 (5.5) |
| NSAID use, n (%) |  |  |  |  |  |  |  |  |  |  |  |
| Use | 1346 (14.0) | 297 (15.4) | 236 (12.3) | 307 (16.0) | 235 (12.2) | 311 (16.2) | 254 (13.2) | 315 (16.4) | 225 (11.7) | 249 (12.9) | 252 (13.1) |
| No use | 7670 (79.8) | 1486 (77.2) | 1585 (82.4) | 1479 (76.9) | 1588 (82.6) | 1499 (77.9) | 1561 (81.2) | 1474 (76.6) | 1572 (81.7) | 1530 (79.5) | 1565 (81.4) |
| Missing | 600 (6.2) | 141 (7.3) | 102 (5.3) | 138 (7.2) | 100 (5.2) | 114 (5.9) | 108 (5.6) | 135 (7.0) | 126 (6.6) | 145 (7.5) | 106 (5.5) |
| Aspirin use, n (%) |  |  |  |  |  |  |  |  |  |  |  |
| Use | 1321 (13.7) | 285 (14.8) | 228 (11.9) | 286 (14.9) | 241 (12.5) | 303 (15.7) | 238 (12.4) | 277 (14.4) | 229 (11.9) | 240 (12.5) | 223 (11.6) |
| No use | 7695 (80.0) | 1498 (77.9) | 1593 (82.8) | 1500 (78.0) | 1582 (82.3) | 1507 (78.3) | 1577 (82.0) | 1512 (78.6) | 1568 (81.5) | 1539 (80.0) | 1594 (82.9) |
| Missing | 600 (6.2) | 141 (7.3) | 102 (5.3) | 138 (7.2) | 100 (5.2) | 114 (5.9) | 108 (5.6) | 135 (7.0) | 126 (6.6) | 145 (7.5) | 106 (5.5) |
| Family history of cardiovascular disease, n (%) |  |  |  |  |  |  |  |  |  |  |  |
| No | 5450 (56.7) | 1186 (61.6) | 1007 (52.4) | 1159 (60.2) | 995 (51.7) | 1142 (59.4) | 1040 (54.1) | 1182 (61.4) | 1002 (52.1) | 1094 (56.9) | 1072 (55.7) |
| Yes | 750 (7.8) | 130 (6.8) | 146 (7.6) | 153 (8.0) | 153 (8.0) | 141 (7.3) | 182 (9.5) | 172 (8.9) | 151 (7.9) | 130 (6.8) | 181 (9.4) |
| Missing | 3416 (35.5) | 608 (31.6) | 770 (40.0) | 612 (31.8) | 775 (40.3) | 641 (33.3) | 701 (36.5) | 570 (29.6) | 770 (40.0) | 700 (36.4) | 670 (34.8) |
| **Family history of diabetes, n (%)** |  |  |  |  |  |  |  |  |  |  |  |
| No | 4544 (47.3) | 972 (50.5) | 832 (43.3) | 978 (50.8) | 837 (43.5) | 947 (49.2) | 908 (47.2) | 996 (51.8) | 856 (44.5) | 896 (46.6) | 940 (48.9) |
| Yes | 1656 (17.2) | 344 (17.9) | 321 (16.7) | 334 (17.4) | 311 (16.2) | 336 (17.5) | 314 (16.3) | 358 (18.6) | 297 (15.4) | 328 (17.0) | 313 (16.3) |
| Missing | 3416 (35.5) | 608 (31.6) | 770 (40.0) | 612 (31.8) | 775 (40.3) | 641 (33.3) | 701 (36.5) | 570 (29.6) | 770 (40.0) | 700 (36.4) | 670 (34.8) |

Data are presented as mean (standard deviation, SD) or median (interquartile range, IQR) for continuous variables and number (percentage) for categorical variables. AHEI, Alternative Healthy Eating Index; AMED, Alternate Mediterranean Diet
Score; BMI, body mass index; hPDI, healthy Plant-based Diet Index; MET-h, metabolic equivalent of task hours; NSAIDs, nonsteroidal anti-inflammatory drugs; Q, quintile; rDII, reversed Dietary Inflammatory Index; rEDIH, reversed Empirical
Dietary Index for Hyperinsulinemia.

### Associations between healthy diet adherence and prevalent MASLD

We defined prevalent MASLD as a self-reported diagnosis (International Classification of Diseases, 11^th^ Revision; ICD-11: DB92/DB92.1) or liver SoS < 1524 m/s (1,579 prevalent MASLD cases; 7,705 without MASLD) (see **Methods** and **Supplementary Figure 3**). Greater adherence to each of the five dietary patterns was inversely associated with prevalent MASLD after adjustment for demographic and lifestyle confounders (Model 2). Multivariable-adjusted odds ratios (ORs) with 95% confidence intervals (CIs) per SD increase in dietary pattern scores ranged from 0.79 (95% CI: 0.73 to 0.84) for the AHEI to 0.88 (95% CI: 0.82 to 0.94) for the rEDIH. These associations were partially attenuated after additional adjustment for BMI (Model 3) and were not materially altered by adjustment for NSAID use and family history of diabetes or CVD (Model 4) (**Figure 2A**).

**Figure 2.**
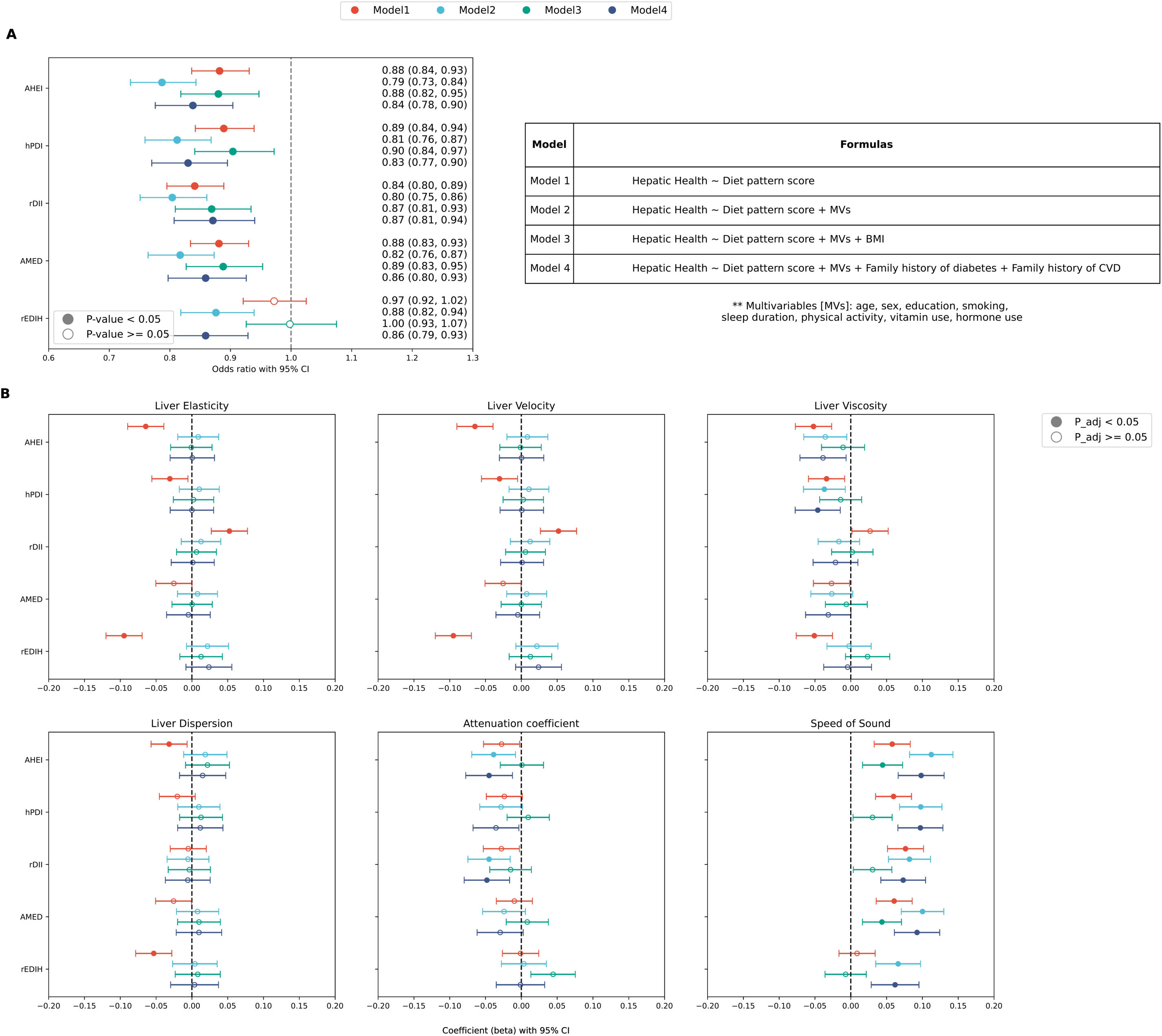
Associations between adherence to different dietary patterns and hepatic health. Forest plots (**A**) show the odds ratios (ORs) with 95% confidence intervals (CIs) of MASLD prevalence for greater adherence to different dietary patterns. The ORs and 95% CIs were calculated by logistic regression models. Based on the hepatic metrics derived from liver ultrasound and 2D-SWE, the associations of healthy diet adherence with six hepatic metrics (liver elasticity, velocity, viscosity, dispersion, attenuation coefficient and speed of sound) are presented in **B**. The β coefficients with 95% CIs were calculated by generalized linear models (GLMs). Four sequential models were fitted for both logistic regression models and GLMs: Model 1,unadjusted; Model 2, with adjustment for age, sex, education level, smoking status, sleep duration, physical activity, vitamin use and hormone use; Model 3, with adjustment for variables in Model 2 plus BMI; Model 4, with adjustment for variables in Model 2 plus NSAID use, family history of diabetes, and family history of CVD. An additional adjustment for alcohol intake was made when hPDI served as the exposure. In **A** and **B**, the solid dots represent associations that remained significant after multiple testing corrections (*FDR*-adjusted *P* < 0.05). AHEI, Alternative Healthy Eating Index; AMED, Alternate Mediterranean Diet Score; BMI, body mass index; CVD, cardiovascular disease; FDR, false discovery rate; hPDI, healthy Plant-based Diet Index; MASLD, Metabolic dysfunction-associated steatotic liver disease; NSAIDs, nonsteroidal anti-inflammatory drugs; rDII, reversed Dietary Inflammatory Index; rEDIH, reversed Empirical Dietary Index for Hyperinsulinemia.

Regarding potential effect modifications, we observed no significant interactions between most dietary pattern scores and sex or BMI category (< 25 vs ≥ 25 kg/m^2^); the only exception was the rDII, for which the association with prevalent MASLD was modified by both sex and BMI (both *P* for interaction = 0.01). The inverse association for rDII was more pronounced in women than in men (OR: 0.73, 95% CI: 0.66 to 0.80 vs 0.88, 95% CI: 0.80 to 0.97) and in participants with BMI ≥ 25 kg/m^2^ than in those with BMI < 25 kg/m^2^ (OR: 0.80, 95% CI: 0.74 to 0.87 vs 0.97, 95% CI: 0.85 to 1.10) (**Supplementary Figure 4A–B**). Although interaction tests for the remaining dietary pattern scores were not statistically significant, inverse associations were generally more pronounced among participants with BMI ≥ 25 kg/m^2^ than among those with BMI < 25 kg/m^2^. Restricted cubic spline models showed no evidence of nonlinear associations between the five dietary pattern scores and prevalent MASLD (**Supplementary Figure 5**), and the results remained similar when scores were analyzed by quintiles (**Supplementary Figure 6**). In a sensitivity analysis restricting MASLD cases to self-reported diagnosis, the AHEI, rEDIH, and hPDI were inversely associated with prevalent MASLD, whereas estimates for the rDII and AMED were directionally consistent but did not reach statistical significance (Model 2, **Supplementary Figure 7**).

### Associations between healthy diet adherence and hepatic metrics from ultrasound

We next examined whether healthy diet adherence is associated with hepatic metrics derived from liver ultrasound, which probably reflect early-stage liver disease prior to overt MASLD. Among 5,989 participants without self-reported MASLD who underwent liver ultrasound, greater adherence to each of the five dietary patterns (per 1-SD increase) was associated with higher liver SoS, but not with the other fibrosis- or inflammation-related metrics, after adjustment for demographic and lifestyle confounders and multiple testing corrections (Model 2, **Figure 2B**). The β coefficients ranged from 0.07 (95% CI: 0.03 to 0.10) for the rEDIH to 0.11 (95% CI: 0.08 to 0.14) for the AHEI (**Supplementary Table 1**). These associations were present in both sexes, with slightly higher estimates in women than in men, and were most pronounced in participants with BMI ≥ 25 kg/m^2^. No associations were observed between dietary pattern scores and other hepatic metrics in sex- or BMI-stratified analyses (**Supplementary Figure 8A–B**). Quintile analysis further indicated a dose-response pattern, with participants in the highest quintile (Q5) showing a statistically significant increase in liver SoS compared with those in the lowest quintile (Q1) (**Supplementary Figure 9**). Sensitivity analyses addressing potential outlier hepatic metric values, either by truncating the values or excluding affected participants, yielded similar conclusions (**Supplementary Figure 10A–B**).

### Associations between healthy diet adherence and overall gut microbial composition

To explore potential links between diet and hepatic health, we examined associations between healthy diet adherence and gut microbial composition, given the proposed role of the gut–liver axis in MASLD pathogenesis^34^. Species belonging to the *Firmicutes* and *Bacteroidetes* phyla accounted for the largest share of the fecal microbial community, consistent with findings from previous population-based cohorts^35–37^ (**Supplementary Figure 11A**). Alpha diversity, measured by Shannon and Simpson indices, differed across quintiles of AHEI, hPDI, AMED, and rEDIH scores (Kruskal–Wallis test, *P* values ranging from 0.0001 to 0.01), but not the rDII (*P* = 0.23 for Shannon index; 0.47 for Simpson index) (**Supplementary Figure 11B**). In multivariable linear regression models, dietary pattern scores combined with age, sex, BMI, smoking status, education level, sleep duration, and physical activity explained 5.25% of Shannon index variance. For individual dietary pattern scores assessed separately, the AHEI accounted for the largest proportion of variance (R^2^, % = 0.42) (**Supplementary Table 2**), consistent with the proportion of microbiome variation explained by diet in prior population-level studies^37,38^. For beta diversity, principal coordinates analysis (PCoA) based on Bray-Curtis dissimilarity showed that the first two coordinates captured 18.65% of variance in species-level composition (PC1: 11.0%; PC2: 7.65%). Permutational multivariate analysis of variance (PERMANOVA; 999 permutations) indicated that all five dietary pattern scores were associated with microbiome beta diversity (*P* = 0.001; **Supplementary Figure 11C**), although no clear visual separation was apparent between the highest (Q5) and lowest (Q1) quintiles for each of the dietary pattern scores. The proportion of variance in microbial composition explained by each dietary score (PERMANOVA, R^2^, %) ranged from 0.31 (rEDIH) to 0.55 (rDII).

### Gut microbial species associated with healthy diet adherence

Beyond overall microbial structure, we examined associations of healthy diet adherence with the abundance of individual microbial species. After quality control, 379 species were retained for analysis (see **Methods**). After adjustment for demographic and lifestyle confounders (Model 2), 197 to 271 species were statistically associated with individual dietary pattern scores, with rEDIH yielding the fewest and hPDI the most associations. The strongest positive association was observed for rDII with *Blautia* sp. AF19 10LB (β = 0.24, 95% CI: 0.22 to 0.27), and the strongest inverse association for rDII with *Clostridium phoceensis* (β = −0.25, 95% CI: −0.27 to −0.22) (Model 2, **Supplementary Table 3**).

We identified 138 microbial species consistently associated with all five dietary pattern scores in the same direction of effect (**Figure 3A**), a conservative intersection criterion requiring each species to independently pass *FDR* correction in each analysis. Most of these species belong to the phyla *Firmicutes*, *Bacteroidetes,* and *Proteobacteria*. Among species inversely associated with healthy diet adherence, the strongest associations were observed for *Clostridium phoceensis, Ruminococcus torques, Flavonifractor plautii*, *Blautia massiliensis*, *Dysosmobacter welbionis*, *Ruthenibacterium lactatiformans*, *Allisonella histaminiformans*, *Phocea massiliensis*, and *Bilophila wadsworthia* (β ranging from −0.07 to −0.25, **Figure 3B**). Among positively associated species, the strongest associations were observed for *Blautia* sp. AF19 10LB, *Clostridium* sp. AF20 17LB, unclassified *Lachnospiraceae* bacterium, unclassified *Bacilli* bacterium, and *Ruminococcus* sp. AF41 9 (βs ranging from 0.03 to 0.24, **Figure 3C**). Adherence to each of the five dietary patterns was also positively associated with the abundance of *Akkermansia muciniphila* (e.g., AHEI-related β: 0.04, 95% CI: 0.02 to 0.07, **Supplementary Table 3**), a species linked to mucosal and metabolic health^39^. Beyond these shared associations, a total of 30 species were exclusively associated with a single dietary pattern score, with rEDIH accounting for the majority (n = 17) (**Supplementary Figure 12** and **Supplementary Table 4**).

**Figure 3.**
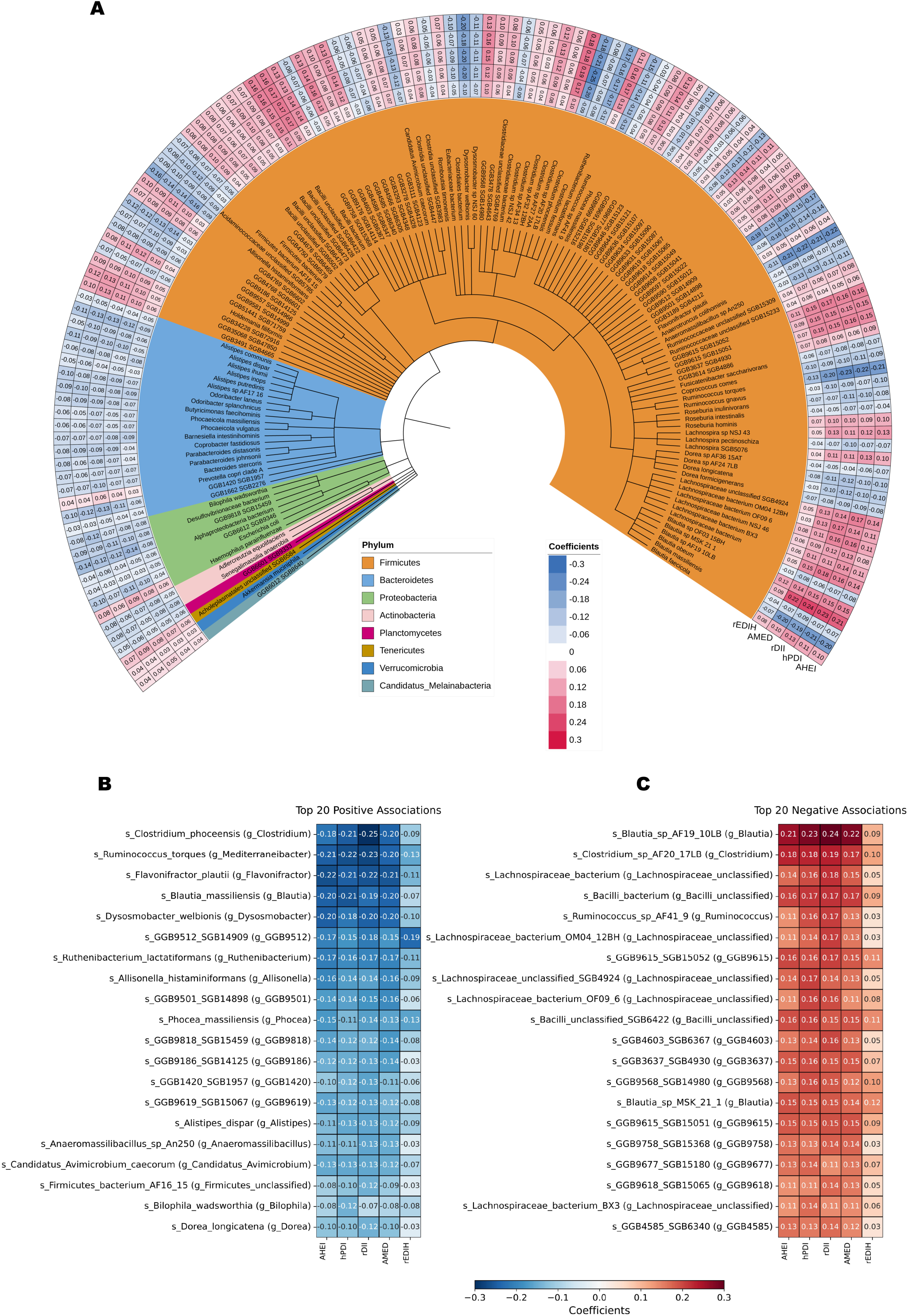
Associations between adherence to healthy dietary patterns and abundance of individual microbial species. The circular heatmap with taxonomic tree shows the consistent microbial species significantly associated with healthy diet adherence (**A**). Each ring of the heatmap indicates one dietary pattern score (AHEI, hPDI, rDII, AMED, and rEDIH). The numbers in the cells of the heatmap indicate the β coefficients (rounded to 2 decimal places) obtained from generalized linear models (GLM). The models considered Z-scores of microbial species abundances (after centered log-ratio [CLR] transformation) as the dependent variables, and different scaled dietary pattern scores as the independent variables, with adjustment for the covariates: age, sex, education level, smoking status, sleep duration, physical activity, vitamin use, and hormone use. When modeling hPDI as the independent variable, alcohol intake was additionally adjusted for. Red colors denote positive associations while blue colors denote negative associations. The strongest (20 positive and 20 negative) associations between adherence to healthy dietary patterns and microbiome abundances are displayed in **B** and **C**. The color scale in **B** and **C** reflects association strength, with red and blue representing positive and negative associations, respectively. All associations shown in the plot passed corrections for multiple testing with a False Discovery Rate (*FDR*)-adjusted *P* < 0.05. AHEI, Alternative Healthy Eating Index; AMED, Alternate Mediterranean Diet Score; FDR, false discovery rate; hPDI, healthy Plant-based Diet Index; rDII, reversed Dietary Inflammatory Index; rEDIH, reversed Empirical Dietary Index for Hyperinsulinemia.

In the sensitivity analyses, associations of healthy diet adherence with the 138 microbial species remained statistically significant for at least one of the five dietary pattern scores after further adjustment for BMI (Model 3) or antibiotic and proton pump inhibitor (PPI) use during the dietary logging period (Model 4) (**Supplementary Table 5**). Of these, 116 species retained statistically significant associations with all five dietary pattern scores across four models (**Supplementary Table 6**).

### Association of diet-related gut microbial species with hepatic outcomes

Of the 138 microbial species consistently associated with all five dietary pattern scores (**Figure 3A**), 27 were also statistically associated with at least one hepatic ultrasound metric in the fully adjusted model. The number of associated species varied across metrics, from 16 (liver SoS) to 1 (liver dispersion) (**Figure 4A**). For prevalent MASLD, 7 species were associated with reduced odds of prevalent MASLD, whereas 13 species (e.g., *Dysosmobacter welbionis, Ruthenibacterium lactatiformans, Flavonifractor plautii, Bilophila wadsworthia*, and *Phocea massiliensis*) were associated with elevated odds of prevalent MASLD after multiple testing corrections (**Figure 4B**). Directional consistency was observed across analyses: species positively associated with liver SoS were also associated with lower odds of prevalent MASLD. Several species inversely associated with liver SoS, including *Clostridium fessum*, *Ruminococcus gnavus*, *Roseburia intestinalis*, and *Parabacteroides distasonis*, showed positive but statistically non-significant point estimates for MASLD prevalence (**Figure 4A–B** and **Supplementary Table 7**). Among the 30 species exclusively associated with a single dietary pattern score, only two were associated with the attenuation coefficient: GGB9705 SGB15224 (β = −0.06, 95% CI: −0.09 to −0.02) and *Clostridium* sp. AM22 11AC (β = 0.08, 95% CI: 0.05 to 0.11) (**Supplementary Figure 13**). Nevertheless, none of those 30 species were statistically associated with prevalent MASLD after correction for multiple testing (**Supplementary Figure 14**).

**Figure 4.**
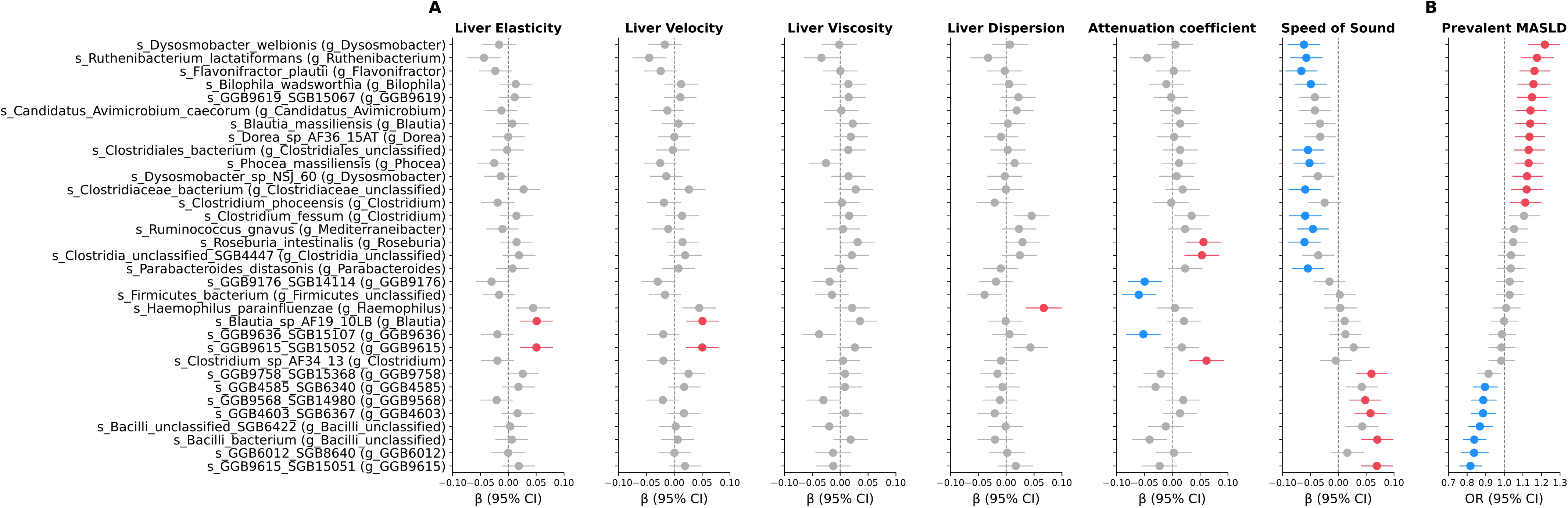
Association of healthy diet adherence-associated gut microbial species and hepatic health. Forest plot (**A**) shows the associations between healthy diet adherence-related microbial species and hepatic metrics derived from liver ultrasound and 2D-SWE, including speed of sound (SoS), attenuation coefficient, liver dispersion, viscosity, velocity, and elasticity. The X-axis displays the β coefficients with 95% confidence intervals (CIs) as error bars for different microbial species. Red colors denote positive associations while blue colors denote negative associations. In **B**, forest plot shows the associations between healthy diet adherence-related microbial species and prevalent MASLD. The X-axis displays the odds ratios (ORs) with 95% CIs as error bars. ORs > 1 denote increased odds of prevalent MASLD (red color), whereas ORs < 1 denote lower odds of prevalent MASLD (blue color). In both **A** and **B**, the Y-axis lists the gut microbial species that were significantly associated with at least one hepatic metric and prevalent MASLD. The statistical models were adjusted for age, sex, BMI, education level, smoking status, sleep duration, physical activity, alcohol intake, vitamin use and hormone use, antibiotic and proton pump inhibitor (PPI) use during the dietary logging period. AHEI, Alternative Healthy Eating Index; AMED, Alternate Mediterranean Diet Score; BMI, body mass index; hPDI, healthy Plant-based Diet Index; MASLD, Metabolic dysfunction-associated steatotic liver disease; NSAIDs, nonsteroidal Anti-inflammatory drugs; rDII, reversed Dietary Inflammatory Index; rEDIH, reversed Empirical Dietary Index for Hyperinsulinemia.

### Gut microbial species as putative mediators in diet–liver associations

To assess whether gut microbial composition may account for a portion of the observed diet–hepatic health associations, we performed cross-sectional mediation analyses focusing on the 16 liver SoS-associated microbial species (**Figure 4A**). All mediation models were adjusted for BMI, given its substantial attenuation of diet–hepatic health associations (Model 3; **Figure 2A–B**). Because the associations of rEDIH with liver SoS and prevalent MASLD were no longer significant after BMI adjustment, mediation analyses were restricted to AHEI, AMED, rDII, and hPDI. Of the 16 liver SoS-associated species, 14 were identified as candidate mediators of the AHEI–SoS and AMED–SoS associations, with *Flavonifractor plautii* showing the largest estimated mediated proportion (∼ 24%), followed by GGB9615 SGB15051 (∼ 22%), *Dysosmobacter welbionis* (∼ 21%), and *Ruthenibacterium lactatiformans* (∼ 19%) (**Figure 5** and **Supplementary Table 8**). For rDII, 11 species were candidate mediators, with GGB9615 SGB15051 (∼ 31%) and *Flavonifractor plautii* (∼ 30%) showing the largest proportions. No species were identified as candidate mediators of the hPDI–SoS association after multiple testing corrections (**Figure 5**). The above candidate mediator species also showed statistical patterns consistent with mediation of the diet–MASLD associations (**Supplementary Figure 15**).

**Figure 5.**
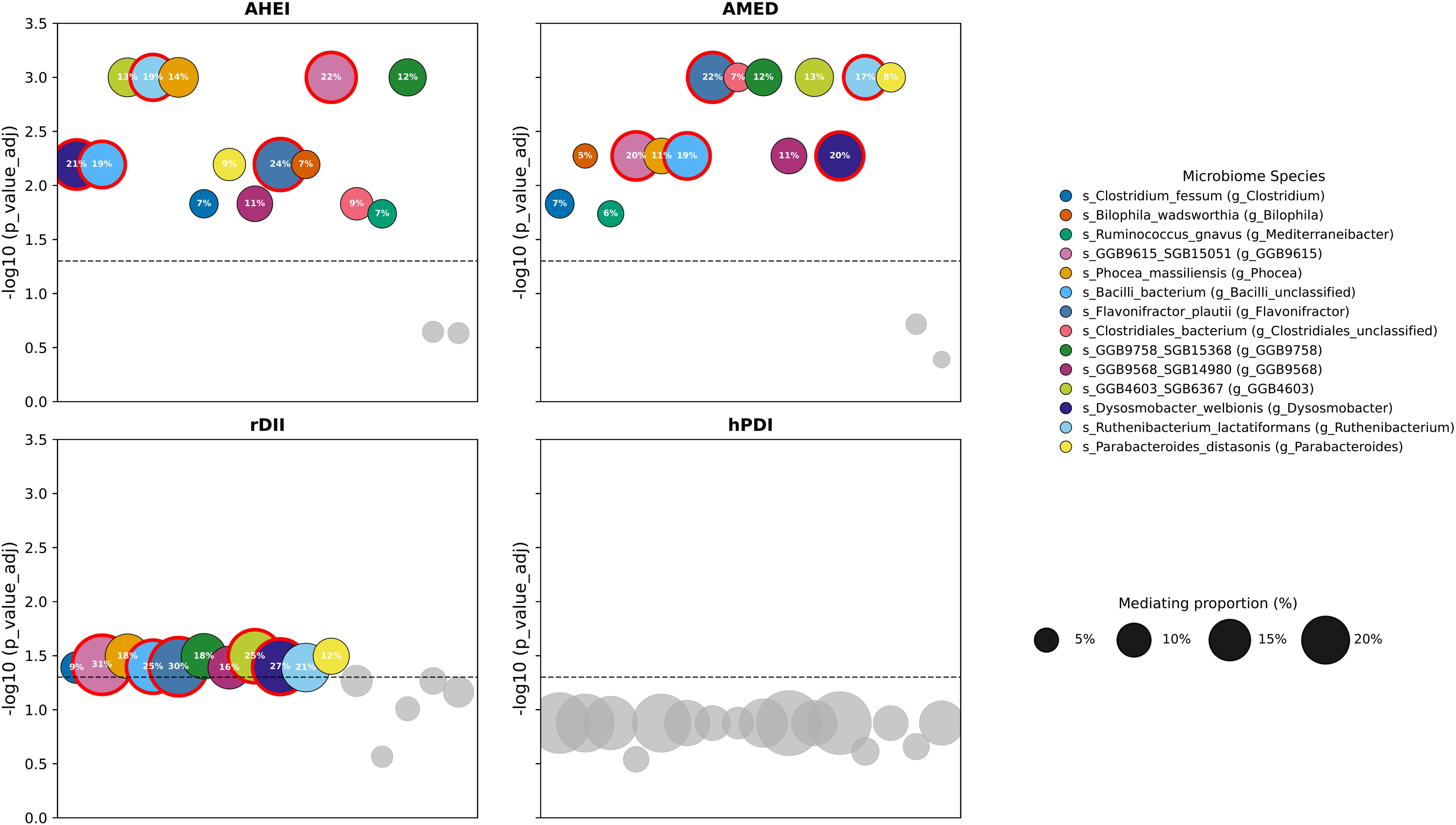
Mediating roles of microbial species between healthy diet adherence and liver speed of sound. Cross-sectional mediation analyses were conducted to examine whether microbial species potentially account for the association between healthy diet adherence and liver SoS. The bubble plots display the mediated proportion for 16 SoS- and healthy diet-associated microbial species in the diet–SoS relationship. The analyses were adjusted for age, sex, BMI, education level, smoking status, sleep duration, physical activity, vitamin use, hormone use, antibiotic and proton pump inhibitor (PPI) use during the dietary logging period. An additional adjustment for alcohol intake was made when hPDI served as the exposure. The *P* values for the mediation analyses were adjusted using Benjamini–Hochberg method. The Y-axis represents the -log10 of the *FDR*-adjusted *P* value for the mediated proportion, with the dashed line denoting the -log10(*FDR*-adjusted *P* value at 0.05). The circle colors denote different microbial species, and grey color represents microbial species with non-significant mediating roles (*FDR-*adjusted *P* value > 0.05). The top five microbial species with the largest mediated proportion are highlighted with red circles. AHEI, Alternative Healthy Eating Index; AMED, Alternate Mediterranean Diet Score; BMI, body mass index; FDR, false discovery rate; hPDI, healthy Plant-based Diet Index; rDII, reversed Dietary Inflammatory Index; SoS, speed of sound.

## DISCUSSION

In this cross-sectional analysis of a deeply phenotyped and population-based cohort, we observed three principal findings. First, adherence to each of the five healthy dietary patterns was inversely associated with MASLD prevalence and positively associated with liver SoS. Second, shotgun metagenomic profiling identified 138 microbial species, the abundance of which was consistently associated with adherence to five correlated but distinct dietary patterns, extending prior 16S rRNA-based observations to species-level resolution^40–43^. Third, mediation analyses identified a set of microbial species, including *Flavonifractor plautii*, *Dysosmobacter welbionis, Ruthenibacterium lactatiformans, Bilophila wadsworthia*, and *Phocea massiliensis*, as candidate intermediaries of the statistical association between diet and liver health.

The inverse association between healthy diet adherence and MASLD prevalence is consistent with prior studies^4,10–17^. Our study extends existing evidence by computing five dietary pattern scores from 2-week continuous dietary logs rather than food frequency questionnaires (FFQs) or 24-hour dietary recalls used in most previous studies^10,14–17,44^, thereby reducing recall bias and measurement error in dietary assessment. Among the scores examined, AHEI showed the strongest inverse association with MASLD prevalence, aligning with its established association with healthy aging^45^. In sensitivity analyses restricting MASLD to self-reported diagnosis, rDII and AMED associations were directionally consistent but did not reach statistical significance, likely due to reduced power from the smaller number of self-reported cases. To our knowledge, no prior studies have examined associations between healthy diet adherence and 2D-SWE-derived hepatic metrics. We therefore assessed six parameters related to steatosis, fibrosis, and inflammation and found that only steatosis-related liver SoS was positively associated with healthy diet adherence. This suggests that the cross-sectional association between healthy diet adherence and hepatic health is primarily detectable through steatosis-related metrics, consistent with prior evidence linking better dietary quality to lower hepatic fat^4,46^. This interpretation is further supported by the inverse correlation between liver SoS and magnetic resonance imaging-derived proton density fat fraction (MRI-PDFF)^47^, positioning SoS as a non-invasive marker of dietary influences on hepatic steatosis in population-based settings.

Beyond the diet–liver association, several of the diet-microbiome associations identified in this study converge with findings from the ZOE PREDICT trial^29,31^, Mediterranean diet intervention trials^26,48^, and observational studies^32,49^. Specifically, *Flavonifractor plautii*, *Ruminococcus torques*, *Ruminococcus gnavus*, *Ruthenibacterium lactatiformans*, *Bilophila wadsworthia,* and *Phocea massiliensis* were each inversely associated with healthy diet adherence, and a Mediterranean diet intervention in overweight and obese participants reduced the abundance of all six species^48^. Among species positively associated with healthy diet adherence, *Blautia* sp. AF19 10LB, *Blautia* sp. MSK 21 1, and multiple unclassified *Lachnospiraceae* bacteria are recognized as butyrate producers, which have been implicated in supporting intestinal barrier integrity and immune homeostasis^50,51^. Their enrichment with healthier diets is corroborated by the ZOE-BIOME intervention trial, in which a prebiotic dietary intervention (rich in plant polyphenols, fiber, and micronutrients) increased the abundance of *Blautia* sp. AF19 10LB and *Lachnospiraceae* bacteria^31^. Consistent with prior observations^29,31^, most species positively associated with healthy diet adherence in the present study remain taxonomically uncharacterized, underscoring a knowledge gap between population-level microbial associations and mechanistic understanding.

The central contribution of this study is the identification of multiple diet-related microbial species that are also associated with hepatic health and partially accounted for the diet–liver association. The gut–liver axis provides a plausible biological framework for these findings.

Previous studies have suggested that species compromising intestinal barrier integrity could increase portal delivery of lipopolysaccharides and other pathogen-associated molecular patterns (PAMPs) to the liver, thereby promoting hepatic inflammation and lipogenesis^34,52^. Aligning with this framework, *Flavonifractor plautii*, *Ruthenibacterium lactatiformans, Bilophila wadsworthia*, *Ruminococcus gnavus*, and *Phocea massiliensis* were each inversely associated with healthy diet adherence yet related to higher MASLD prevalence and lower liver SoS. For several of these species, existing studies have offered direct links to hepatic health. *Bilophila wadsworthia* has been shown to synergize with high-fat diets to promote intestinal barrier dysfunction and hepatic steatosis in murine models and has been positively associated with red meat intake^32^ and adverse cardiometabolic outcomes^53,54^. *Ruminococcus gnavus*, a mucin-degrading species that compromises the intestinal mucus layer, has been consistently enriched in hepatic steatosis and fibrosis^55,56^, and both *Ruminococcus gnavus* and *Phocea massiliensis* showed positive associations with fatty liver index in a Finnish population^57^. For other species, evidence linking them directly to hepatic health remains limited and studies have implicated them in related cardiometabolic pathways. For example, *Flavonifractor plautii* abundance was associated with lower insulin sensitivity, reduced disposition index, and higher dyslipidemia prevalence^58^, and with elevated cardiovascular risk alongside *Ruthenibacterium lactatiformans*^59^. A meta-analysis from the MicroCardio Consortium further showed that *Flavonifractor plautii*, *Bilophila wadsworthia*, and *Ruminococcus gnavus* were all positively associated with T2D^60^; in the IMI-DIRECT prospective study, increased *Ruthenibacterium lactatiformans* abundance paralleled metabolic deterioration in individuals with prediabetes was also reported^61^. Collectively, our findings suggest that healthy diet-associated gut microbial species are closely linked to host metabolic health. Given the shared pathophysiology between cardiometabolic dysfunction and MASLD, the observed associations provide biological plausibility for a potential role of species not yet directly implicated in hepatic health.

Regarding the heterogeneity in the mediating roles of gut microbial species across diet–hepatic health associations, the absence of significant microbial mediators for the hPDI–SoS association probably reflects the distinct dietary information captured by this index. The association between plant-based dietary patterns and liver steatosis could involve other non-microbial pathways such as plant-derived phytochemicals and fiber^62^. Nonetheless, when prevalent MASLD was the outcome, gut microbial species showed more consistent mediating roles across dietary patterns. This divergence plausibly indicates the differing nature of the two hepatic metrics. Liver SoS, as a continuous measure, captures subtle and pathway-specific variation in how different diets impact liver steatosis. By contrast, prevalent MASLD, a binary clinical outcome, reflects converging metabolic dysfunction, which tends to yield more uniform microbial mediation across dietary patterns. However, this interpretation remains speculative and warrant investigation in the further studies.

An intriguing finding in this study concerns *Dysosmobacter welbionis*, which illustrates the complexity of interpreting population-level microbial associations. Consistent with findings from other cohorts^31,63,64^, this species was inversely associated with healthy diet adherence in our study. Moreover, its higher abundance was associated with both lower liver SOS and higher odds of prevalent MASLD, indicating a consistent link with poor hepatic health. Yet a contrasting picture has emerged from clinical and experimental studies: in humans with obesity and T2D, *Dysosmobacter welbionis* abundance correlated negatively with BMI, fasting glucose, and hepatic aminotransferase levels^65,66^, and supplementation with *Dysosmobacter welbionis* J115^T^ improved glycemia and hepatic steatosis in high-fat-diet-fed mice^67^. This paradox likely indicates context-dependent functionality of this species. Possible explanations are that *Dysosmobacter welbionis* proliferates in the gut environment of metabolically compromised hosts where substrates favoring their growth are more abundant, or that it could occupy an ecological niche displaced by the broader microbial restructuring that accompanies a healthy diet, independent of its intrinsic metabolic effects.

This study has several strengths, including a large cohort with concurrent dietary logs, gut metagenomics, and quantitative liver ultrasound data; comprehensive adjustment for demographic, lifestyle, and medication-related confounders; and consistency of diet–gut–liver associations across five dietary pattern scores. However, several limitations should be noted. First, the cross-sectional design precludes the establishment of temporal relations, and reverse causation remains a concern. For example, MASLD can alter bile acid metabolism and reshape the gut microbiome, and awareness of a liver disease diagnosis may influence dietary behavior. Accordingly, our mediation results should be interpreted as statistical decompositions rather than evidence of causal mediation, as causal inference would require longitudinal data with repeated dietary, metagenomic, and hepatic imaging assessments. Second, residual confounding cannot be excluded. For example, we lacked information on fasting duration before fecal sampling and before liver ultrasound measurement. Third, although 2-week dietary logs improve upon single-time-point food assessment, they capture short-term rather than long-term habitual intake, and some degree of dietary misclassification remains possible. Additionally, certain nutrients required for the original dietary pattern score algorithm were not available in the dietary logs, which could attenuate the strength of observed associations. Fourth, our cohort comprised generally healthy Israeli volunteers with relatively high educational attainment, which may limit generalizability to other populations and clinical settings.

## CONCLUSION

In summary, this study identified associations linking healthy diet adherence, specific gut microbial species, and hepatic health in a large population-based cohort. Several microbial species (e.g., *Flavonifractor plautii*, *Dysosmobacter welbionis*, *Ruthenibacterium lactatiformans, Bilophila wadsworthia*, and *Phocea massiliensis*) partially accounted for diet–liver associations after conditioning on BMI, nominating them as potential targets for microbiome-directed strategies in MASLD prevention, particularly for individuals who find it difficult to adhere to healthy dietary patterns. These findings motivate future interventional studies to evaluate whether modulation of specific gut microbial species mediates the beneficial effects of healthy dietary patterns on hepatic health.

## METHODS

### Study population

This cross-sectional study used data from the HPP, an ongoing large-scale cohort and biobank of healthy adults with deep multi-omics profiling, including metabolomics and gut metagenomics, described in detail previously^33^. Participants aged 40–70 years at baseline were enrolled through voluntary self-referral and a screening survey beginning October 28, 2018, with the aim of identifying novel diagnostic biomarkers and therapeutic targets. Follow-up visits are scheduled biennially over 25 years, with annual questionnaires and comprehensive profiling at the clinical testing center (CTC). CTC assessments include anthropometric measurements, vital signs (e.g., blood pressure), complete blood count, ankle-brachial index, and imaging studies (e.g., liver and carotid ultrasound). During the two weeks following each CTC visit, participants undergo continuous glucose monitoring with concurrent logging of meals, medications, and exercise via a proprietary application, as well as sleep monitoring with a home sleep apnea test device for three nights. At the time of analysis, only Israelis were recruited. Most participants were of European (Ashkenazi) Jewish descent, were free of major chronic diseases, and had high educational attainment.

### Dietary assessment and dietary pattern scores

Participants in the HPP cohort were instructed to log their food intake in real time over two weeks using a designated mobile app (“Project 10K app”), which has been used in previous studies^68,69^. Data from dietary logs correlated well with serum metabolites known to be diet-related, supporting the validity of this assessment method^70^. At the baseline visit, 9,616 participants had complete dietary log data. We focused on five dietary patterns commonly investigated in epidemiological studies of cardiometabolic diseases and derived the corresponding scores following the methods described in previous studies^71,72^. Specifically, we computed three guideline-recommended indices: the Alternative Healthy Eating Index (AHEI), healthy Plant-based Diet Index (hPDI), and Alternate Mediterranean Diet Score (AMED), as well as two mechanism-based indices, the reversed Dietary Inflammatory Index (rDII) and reversed Empirical Dietary Index for Hyperinsulinemia (rEDIH), using data collected via the logging app (see **Supplementary Methods**).

### Ascertainment of prevalent MASLD

As participants in the HPP cohort are relatively healthy at baseline and follow-up is ongoing, we anticipated that only a limited number of incident MASLD cases will occur. Therefore, we identified prevalent MASLD cases through self-reported diagnosis (ICD-11: DB92/DB92.1) and a liver SoS < 1524 m/s measured by ultrasound^73^. As a sensitivity analysis, we re-ran the statistical models using participants with self-reported MASLD diagnosis as cases.

### Measurement of hepatic health metrics

Using the Supersonic Aixplorer MACH 30 ultrasound system (Hologic, USA) with 2D shear wave elastography, we obtained six hepatic measurements: liver elasticity, viscosity, attenuation coefficient, velocity, dispersion, and liver speed of sound^74^. These parameters capture complementary aspects of liver pathology, including fibrosis (liver elasticity [unit: kPa] and velocity [unit: m/s]), inflammation (liver viscosity [unit: Pa.s] and dispersion [unit: (m/s)/kHZ]), and steatosis (attenuation coefficient [unit: dB/cm/MHz] and speed of sound [unit: m/s]), enabling comprehensive, non-invasive assessment of liver health. Specifically, lower speed of sound (m/s) is associated with higher hepatic fat content^75,76^. Liver elasticity is measured in kPa and serves as an indicator of fibrosis, with higher values corresponding to stiffness and fibrosis^77,78^. The detailed measurement protocol was previously reported^79^ (available at: https://knowledgebase.pheno.ai/). At the time of recruitment, 6,738 participants had complete hepatic metrics data.

### Microbiome sample collection and preprocessing

Sample collection, DNA extraction, and sequencing were described previously in detail^80^. Briefly, participants collected stool samples at home with a standardized kit (OMNIgene-GUT OMR-200, DNA Genotek) and transferred them at room temperature to the clinical testing center at the Weizmann Institute of Science, where samples were documented and stored at - 20 °C until processing. Metagenomic DNA was extracted by the PowerMag Microbial DNA Isolation Kit (MO BIO Laboratories, 27200-4). Sequencing libraries were prepared by the NEBNext Ultra II DNA Library Prep Kit for Illumina (New England Biolabs, E7775) with Illumina unique dual indexes (IDT-Syntezza Bioscience) and sequenced on a NovaSeq platform (Illumina) using 100-bp single-end reads, targeting 10 million reads per sample. DNA purification, library preparation, and sequencing were performed in batches of 384 samples, each including a mock microbial community (ZymoBIOMICS Gut Microbiome Standard, D6331) for quality control.

### Metagenomic read mapping and abundance profiling

Sequencing reads were quality-filtered, adapter-trimmed, and host-depleted, then mapped to a genome reference set for taxonomic classification and abundance estimation using the algorithm described in Leviatan *et al*.^81^ Quality control was performed at both the species and sample levels. First, species present in fewer than 10% of samples were removed, resulting in 388 species remaining out of 2,088. Next, species with a mean relative abundance below 0.01% were excluded, leaving 379 species. Sample-level filtering then removed samples with more than 90% missing values across all retained species, resulting in 9,075 individuals remaining out of 9,087. Missing values were imputed using half of the minimum observed abundance for each species, under the assumption that missingness reflects values below the detection limit. The imputed values across all species ranged from 0.005% to 0.3%, with a median of 0.02%. To assess the overall composition of the gut microbiome, we computed diversity metrics prior to centered log-ratio (CLR) transformation: (1) alpha diversity was measured by the Shannon index and the Simpson index, capturing species richness and evenness within each sample; (2) beta diversity was calculated with the Bray-Curtis dissimilarity index, measuring compositional dissimilarity between samples. Differences in beta diversity were tested by PERMANOVA per dietary pattern and beta diversity was visualized through principal coordinates analysis (PCoA). Before statistical analysis, the relative abundance of all species was subjected to the CLR transformation to address the compositional nature of the data, followed by Z-score standardization.

### Assessment of covariates and potential confounders

Potential confounders were selected based on a previous study^71^. Height and weight were measured at baseline, from which BMI was calculated. Information on age, sex, education level, smoking status (current, former or never), physical activity (reported as metabolic equivalent of task hours per day; MET-h/day) and sleep duration (hours/day) was self-reported as part of a pre-baseline visit online questionnaire. Medication use was ascertained by self-reports relating to whether the medication was taken between the study visits. Medication records were coded into Anatomical Therapeutic Chemical (ATC) codes based on the World Health Organization Collaborating Center for Drug Statistics Methodology. For example, vitamin use was defined based on ATC codes starting with A11*; hormone use was based on ATC codes starting with G03*, H01*, H02*, H03*, H04* and H05*; aspirin and other NSAID use was based on ATC codes starting with B01*, M01* and N02*. Medical conditions were ascertained upon enrollment through an online medical questionnaire and through interviews conducted by trained researchers at the assessment center, which also captured approximate dates of onset. Medical diagnoses were linked to ICD-11 codes.

#### Statistical analysis

All analyses were conducted with Python (version 3.11.5) in Jupyter Notebook. Figures were generated using matplotlib (version 3.8.4), seaborn (version 0.13.2), and the *iTOL* software^82^.

Descriptive characteristics were summarized as mean (SD), median (IQR), or number (percentage) for all participants and by quintiles of energy-adjusted dietary pattern scores. All dietary pattern scores were truncated at the 0.5^th^ and 99.5^th^ percentiles to minimize the influence of outliers. Correlations between these scores were assessed with Spearman’s coefficients. Associations of dietary pattern scores with prevalent MASLD and continuous ultrasound-derived hepatic metrics were estimated by logistic and generalized linear models (GLM), respectively. Dietary pattern scores were standardized (mean = 0, SD = 1) to enable cross-pattern comparison of effect sizes. Results were reported as ORs with 95% CIs for logistic models and as β coefficients with 95% CIs for GLM. For hepatic metric analyses, individuals with self-reported MASLD at baseline were excluded to reduce reverse association bias. All hepatic metrics were rank-based inverse-normal transformed.

##### Model specifications

Four sequential models were fitted: Model 1, unadjusted; Model 2, adjusted for age, sex, education level, smoking status, sleep duration, physical activity, vitamin use, and hormone use; Model 3, Model 2 plus BMI; Model 4, Model 2 plus NSAID use, family history of diabetes, and family history of CVD. Because AHEI, AMED, rDII, and rEDIH incorporate alcohol intake as a scored component, alcohol was not adjusted for separately in these models; for hPDI, which does not include alcohol, models were additionally adjusted for alcohol intake (g/day). Model 2 was designated as the primary analysis, and Models 3 and 4 as sensitivity analyses to avoid potential overadjustment for mediators. All analyses used complete cases without imputation, and participants with missing covariate data were excluded from the corresponding models.

##### Nonlinearity and stratification analysis

Nonlinearity in the association between dietary pattern scores (continuous) and prevalent MASLD was assessed using restricted cubic spline models with 3 knots at the 10^th^, 50^th^, and 90^th^ percentiles of dietary scores, adjusted for Model 2 covariates, and tested by likelihood ratio comparison of nested models with and without spline terms. Dietary pattern scores were also categorized by quintiles, with the lowest quintile as reference, and Model 2 was refitted. Interactions between dietary pattern scores (continuous) and sex or BMI category (BMI ≥ 25 and < 25 kg/m^2^) on prevalent MASLD were tested using Wald tests for cross-product terms, with *P* < 0.05 considered significant. Stratified analyses were conducted across sex and BMI strata, adjusted for Model 2 covariates.

##### Sensitivity analyses

The diet–MASLD associations were re-estimated using self-reported MASLD diagnosis as the case definition. For the dietary pattern-hepatic metric associations, to assess the influence of extreme ultrasound-derived values, models were repeated after (1) truncating hepatic metric values at the 1^st^ and 99^th^ percentiles and (2) excluding individuals with liver SoS values exceeding the 99^th^ percentile.

##### Microbiome-wide association and cross-sectional mediation analyses

Microbiome-wide associations with dietary pattern scores were evaluated by first comparing alpha and beta diversity across dietary pattern score quintiles. At the species level, GLMs were fitted with dietary pattern scores as independent variables and CLR-transformed, standardized gut species abundances as dependent variables, using the four sequential models with Model 2 as primary. To estimate associations between diet-related microbial features and liver steatosis, we employed GLMs for continuous metrics and logistic regression for prevalent MASLD. The models were adjusted for age, sex, BMI, education level, smoking status, sleep duration, physical activity, vitamin use, hormone use, alcohol intake (for hPDI only), and antibiotic and PPI use during the dietary logging period, which were selected because antibiotics, PPI use, BMI, and alcohol intake are established confounders of gut microbiome composition. Cross-sectional mediation analysis was performed to assess whether diet-related gut microbiota statistically mediated associations between habitual diet and hepatic health. Participants with missing dependent, independent, or mediator variables were excluded. Each mediation model was adjusted for the microbiome-related covariates. The mediated proportion was calculated as the indirect effect divided by the total effect, with 95% CIs derived via bootstrapping (1,000 iterations) with statsmodels (version 0.14.6) in Python.

##### Multiple testing corrections

For analyses involving multiple comparisons, *P* values were corrected using the Benjamini–Hochberg procedure, with *FDR*-adjusted *P* < 0.05 considered statistically significant. *FDR* correction was applied separately within pre-defined blocks: diet–hepatic metrics (per dietary score), diet–microbiota (species level; 379 tests per score), microbiota–hepatic outcomes, and mediation analysis.

## Data Availability

The individual user data in this study are part of the Human Phenotype Project and are accessible to researchers from universities and other research institutions at https://humanphenotypeproject.org/data-access.

## ACKNOWLEDGEMENTS

We thank all the investigators, the staff, and the participants of the Human Phenotype Project (HPP) cohort for their contributions. This study was conducted using data from the HPP study on behalf of Pheno.AI. Pheno.AI had no role in study design, data analysis, or the decision to submit this manuscript.

## ETHICS

The Human Phenotype Project is conducted according to the principles of the Declaration of Helsinki and was approved by the Institutional Review Board of the Weizmann Institute of Science (protocol no. 964-1; IRB approval number: 1719-1). All participants were fully informed of the study content and provided written informed consent before joining the study.

## FUNDING

R.L-G is supported by the JPI HDHL NUTRIMMUNE DIYUFOOD project. K.D. is supported by the China Scholarship Council (No. 202206210140). The funders had no role in the design, conduct, analysis, or reporting of this study.

## DATA AND CODE AVAILABILITY

The individual user data in this study are part of the Human Phenotype Project and are accessible to researchers from universities and other research institutions at https://humanphenotypeproject.org/data-access. Bona fide researchers should contact to obtain instructions about how to access the data. Computational analysis was performed in Python using the following packages: NumPy and Pandas for data processing, matplotlib and seaborn for creating the figures. The Python code for the analyses in this study is available by the link: https://github.com/Keyong-bio/Microbiome_project.

## COMPETING INTERESTS

E.S. is a paid consultant of Pheno.AI Ltd, a biomedical data science company from Tel Aviv, Israel. All other authors declare no competing interests.

## AUTHOR CONTRIBUTIONS

K.D. and R.L.-G. conceptualized the project. K.D. led the data analysis, interpretation of results, and data visualization, and drafted the manuscript. Q.D, A.G, Z.Z, R.M, A.H.V, F.R.R, G.Z, E.S, and R.L.-G. revised the manuscript. K.D and R.L-G are the guarantors of this work and have full access to all the data in this study. K.D and R.L-G take responsibility for the integrity of the data and the accuracy of the data analysis.

## PRIOR PRESENTATION

A non-peer-reviewed version of this article was posted on the medRxiv preprint server.

## Notes

### Competing Interest Statement

E.S. is a paid consultant of Pheno.AI Ltd. Pheno.AI had no role in study design, data analysis, or the decision to submit this manuscript. All other authors declare no competing interests.

### Author Declarations

The Human Phenotype Project is conducted according to the principles of the Declaration of Helsinki and was approved by the Institutional Review Board of the Weizmann Institute of Science (protocol no. 964-1; IRB approval number: 1719-1).

