## Supplementary Methods and Figures for "Decoding the diet–gut–liver axis: links between dietary pattern adherence, gut microbiome, and hepatic health"

**Running title**: Diet, gut microbiota, and hepatic health

Keyong Deng^1^, Quinten R. Ducarmon^2^, Anastasia Godneva^3,4^, Zheqing Zhang^5^, Astrid van Hylckama Vlieg^1^, Frits R. Rosendaal^1^, Georg Zeller^2^, Eran Segal^3,6^, Ruifang Li-Gao^1^ on behalf of DIYUFOOD consortium

**Affiliations:**

1. Department of Clinical Epidemiology, Leiden University Medical Center, Leiden, the Netherlands
2. Leiden University Center for Infectious Diseases (LUCID), Leiden University Medical Center, Leiden, The Netherlands
3. Department of Computer Science and Applied Mathematics, Weizmann Institute of Science, Rehovot, Israel
4. Department of Molecular Cell Biology, Weizmann Institute of Science, Rehovot, Israel
5. Department of Nutrition and Food Hygiene, Guangdong Provincial Key Laboratory of Tropical Disease Research, School of Public Health, Southern Medical University, Guangdong, China
6. Mohamed bin Zayed University of Artificial Intelligence, Abu Dhabi, UAE

SUPPLEMENTARY MATERIALS

### Dietary logging information and dietary pattern calculations

Dietary intake was self-reported via designated mobile app (“Project 10K app”) in the 2-week period. This app, specifically developed for the study, included more than 7,000 food items with full nutritional values based on Israeli Ministry of Health database, which has been presented in previous studies^1,2^. Participants selected each food item from the database, noting its weight and portion size, and logged it into their user profile in the mobile app. Logging data underwent a quality control process, such as removing items with implausible weights and timing (e.g., many meals logged in a short period). Nutrients were calculated using the United State Department of Agriculture (USDA) Databases. Based on the reported nutrients and food intake, five dietary pattern scores were computed, including Alternate Mediterranean Diet Score (AMED)^3^, Alternative Healthy Eating Index (AHEI)^4^, healthful Plant-Based Diet Index (hPDI)^5^, Empirical Dietary Index for Hyperinsulinemia (EDIH)^6^, and Dietary Inflammatory Index (DII)^7^. The DII and EDIH were empirically derived using biomarkers of chronic inflammation and hyperinsulinemia, respectively. To harmonize all five dietary pattern scores, the DII and EDIH were reversed so that higher values denote greater adherence to healthy dietary patterns.

### Alternate Mediterranean Diet (AMED) score

The AMED score comprised nine components: fruits, vegetables (excluding potatoes), whole grains, nuts, legumes, fish, red and processed meat, the ratio of monounsaturated fat to saturate fat, and alcohol^3^. For most components, participants with intakes above the population median received 1 point; those below received 0. However, we applied one point where reported red and processed meat consumption was less than the population median. Alcohol intake was scored based on sex-specific median values, with 1 point assigned for intake of 5–15 g/d in women and 10–25 g/d in men. The detailed food components used in the Human Phenotype Project (HPP) cohort are shown in **Supplementary Table** **9**.

### Alternative Healthy Eating Index (AHEI)

The AHEI was calculated based on the intake of 11 food and nutrient groups: vegetables, fruits, whole grains, sugar-sweetened beverages and fruit juice, nuts and legumes, red and processed meat, alcohol, sodium, trans fats, long-chain fatty acids, and polyunsaturated fatty acids^4^. Scores ranged from 0 to 10 points per group, proportional to daily intake. Sex-specific criteria were applied to score alcohol intake. The detailed food components used for calculation in the HPP cohort are shown in **Supplementary Table 10**.

### Healthy Plant-based Diet Index (hPDI)

The hPDI categorized 18 food groups into three types: healthy (e.g., whole grains, fruits, vegetables, nuts, legumes, vegetable oils, and tea/coffee); less healthy (e.g., fruit juice, refined grains, potatoes, sugar-sweetened beverages, and sweets/desserts); and animal-based food (e.g., animal fat, dairy, eggs, fish/seafood, meat, miscellaneous animal-based foods)^5^. Quintiles of food group intake (g/day) were used to assign points: 5 pints for the highest quintile of healthy food intake and the lowest quintile of less healthy or animal-based food intake, with 1 point assigned. **Supplementary Table 11** shows the list of food categories included in the HPP cohort.

### Empirical Dietary Index for Hyperinsulinemia (EDIH)

The EDIH was based on 18 food groups related with hyperinsulinemia: red meat, low-energy beverages, cream soup, processed meat, margarine, poultry, butter, potato fries, fish, high-energy beverages, tomato, low-fat dairy, eggs, wine, coffee, fruit, high-fat dairy, vegetables^6^. Food groups were analyzed as grams per day. The EDIH score was calculated as the weighted sum of food-specific intake scores. **Supplementary Table 12** shows the detailed weights for each food component that are available in HPP cohort.

### Dietary Inflammation Index (DII)

Shivappa et al. derived food specific inflammatory effect scores based on 1,934 studies that examined dietary effects on inflammatory biomarkers, including interleukin-1β, interleukin-4, interleukin-10, tumor necrosis factor-alpha, interleukin-6, and C-reactive protein^7^. The DII quantifies dietary inflammatory potential using various food categories. For food groups, the representative world database provides a mean and standard deviation for consumption of each food component in the global population. Food intake of individual was expressed as a relative to the standard global mean as a Z-score: (diet intake – standard global mean) / global SD. To minimize the effect of right-skewing, this value is converted to a percentile score. To further achieve a symmetrical distribution with values centered on 0 and bounded between -1 (maximally anti-inflammatory) and 1 (maximally pro-inflammatory), each percentile score is doubled and then subtract 1. The centered percentile value for each dietary component is multiplied by its respective “overall dietary component-specific inflammatory effect score” to obtain the dietary component-specific DII score. All the dietary component-specific inflammatory effect scores are summed up to indicate overall DII scores for an individual. **Supplementary Table 13** shows the list of food categories included in the HPP cohort.

### Supplementary Figure 1. Distribution of five energy-intake adjusted dietary pattern scores.

A higher dietary pattern score indicates greater adherence to a healthy diet. AHEI, Alternative Healthy Eating Index; AMED, Alternate Mediterranean Diet Score; hPDI, healthy Plant-based Diet Index; rDII, reversed Dietary Inflammatory Index; rEDIH, reversed Empirical Dietary Index for Hyperinsulinemia.

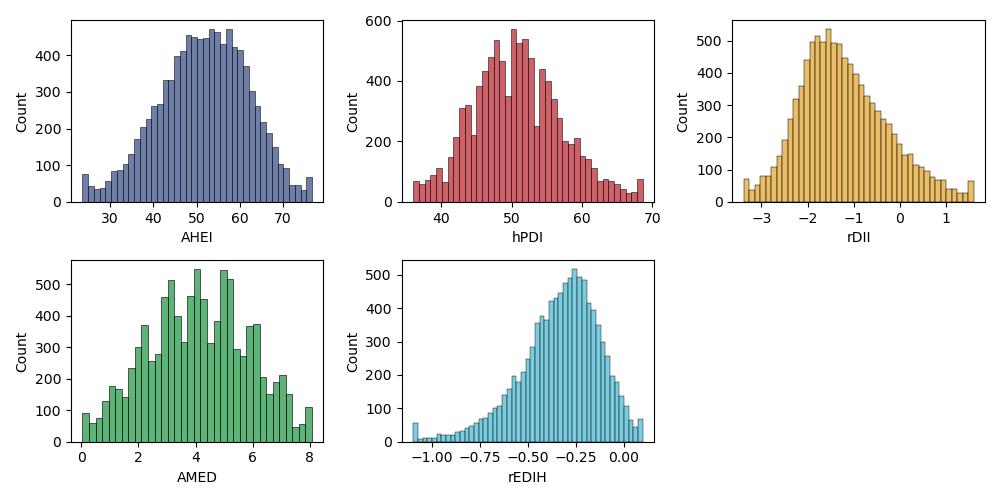

### **Supplementary Figure 2. Spearman correlations between five energy-intake adjusted dietary pattern scores.**

AHEI, Alternative Healthy Eating Index; AMED, Alternate Mediterranean Diet Score; hPDI, healthy Plant-based Diet Index; rDII, reversed Dietary Inflammatory Index; rEDIH, reversed Empirical Dietary Index for Hyperinsulinemia.

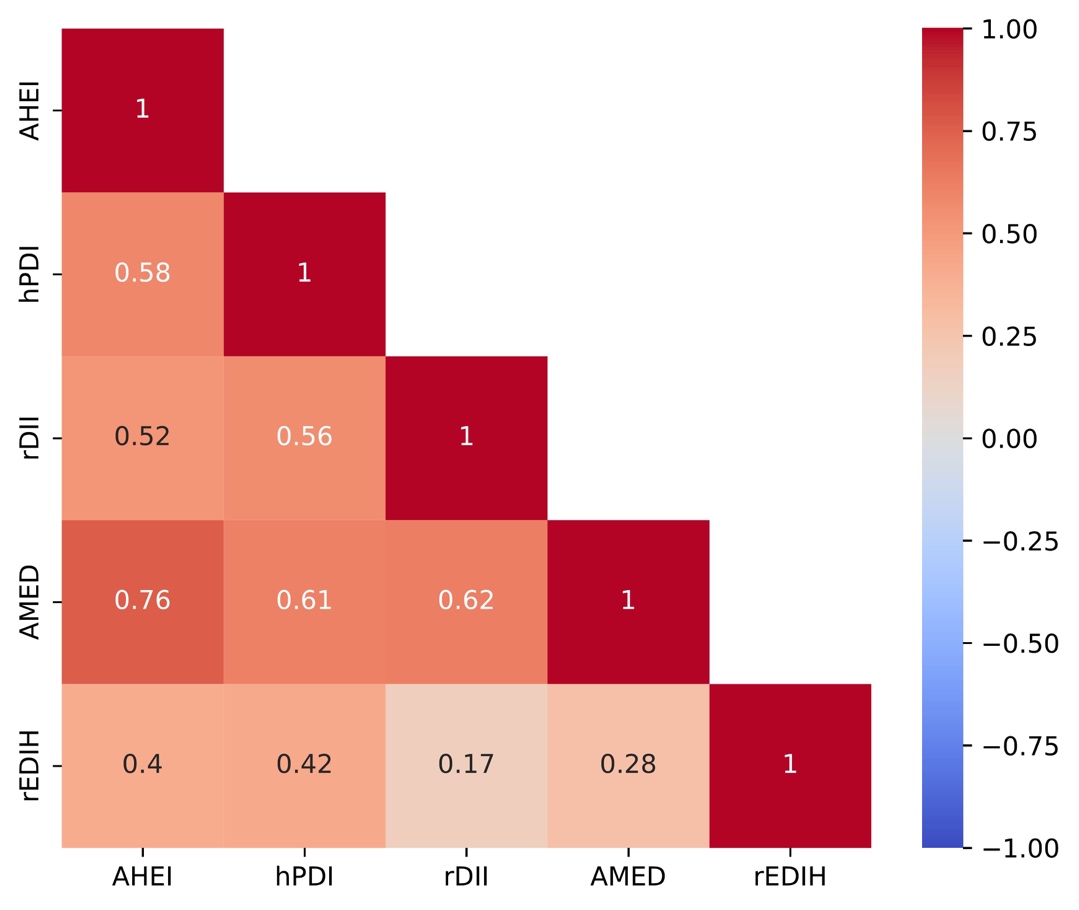

### Supplementary Figure 3. Flowchart of participants selection in the primary analysis.

The flow chart illustrates the exclusion criteria and data integration steps. Initially, 9,616 participants remained after excluding those without dietary logging data. **A** shows the subsequent selection based on available metagenomic and liver ultrasound (2D-SWE) measurements. **B** represents the integrative cohort (n = 5,338) with complete data across all three domains: diet, microbiome, and liver health.

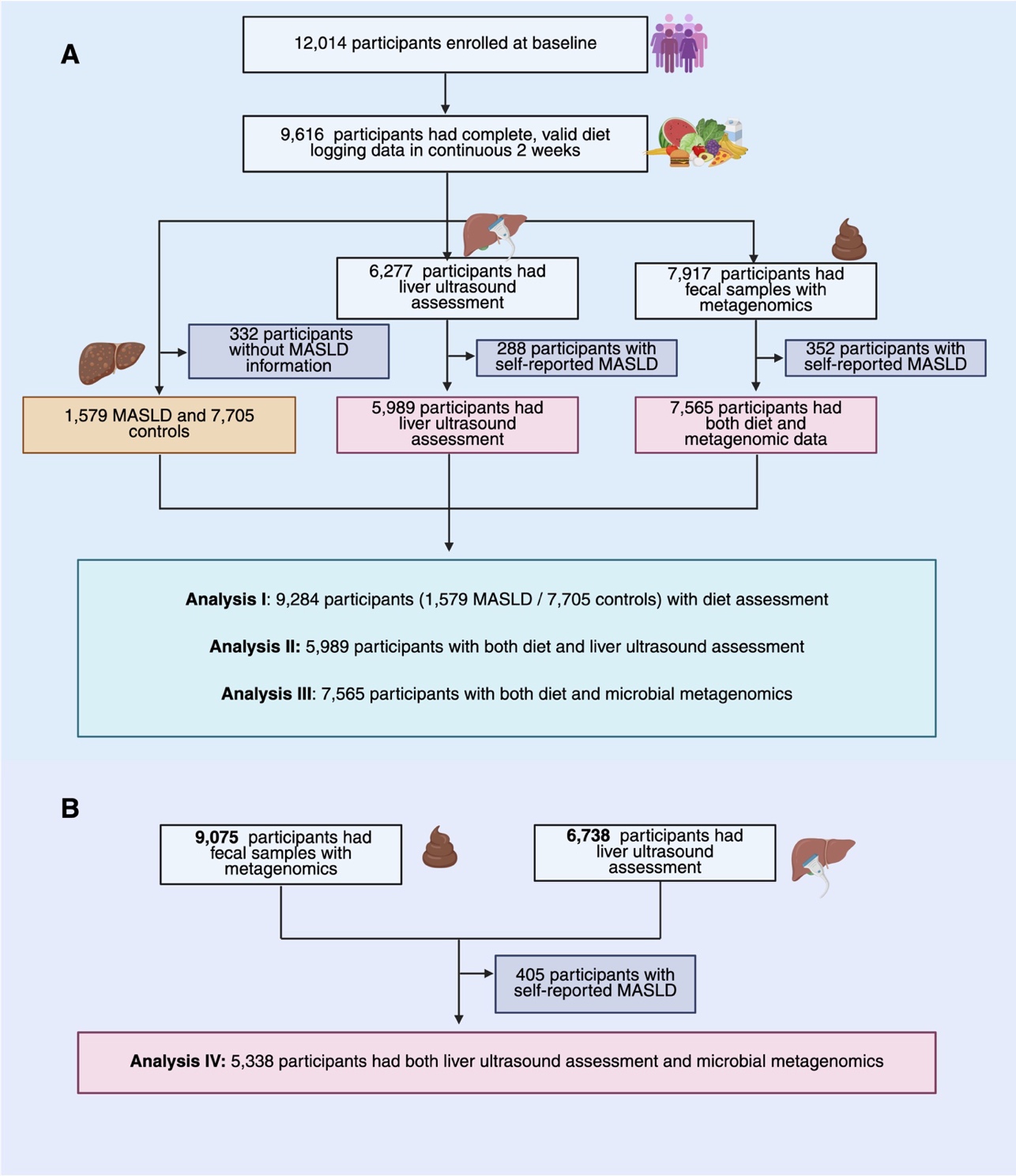

### Supplementary Figure 4. Associations between adherence to healthy dietary patterns and prevalent MASLD, stratified by sex and BMI.

Forest plots show multivariable-adjusted odds ratios (ORs) with 95% confidence intervals (CIs) per SD increase in dietary pattern scores for the association with prevalent MASLD, stratified by sex (**A**) and BMI category (<25 vs. ≥ 25 kg/m²; **B**). In **A**, models were adjusted for age, education level, smoking status, sleep duration, physical activity, vitamin use, and hormone use. In **B**, models were adjusted for the same covariates plus sex. For hPDI, models were additionally adjusted for alcohol intake. The solid dots indicate statistically significant associations (*P* < 0.05); the hollow points indicate non-significant association. Asterisks denote dietary patterns for which a statistically significant interaction (*P*_interaction_ < 0.05) was observed with the corresponding stratification variable. AHEI, Alternative Healthy Eating Index; AMED, Alternate Mediterranean Diet Score; BMI, body mass index; hPDI, healthy Plant-based Diet Index; MASLD, metabolic dysfunction-associated steatotic liver disease; rDII, reversed Dietary Inflammatory Index; rEDIH, reversed Empirical Dietary Index for Hyperinsulinemia.

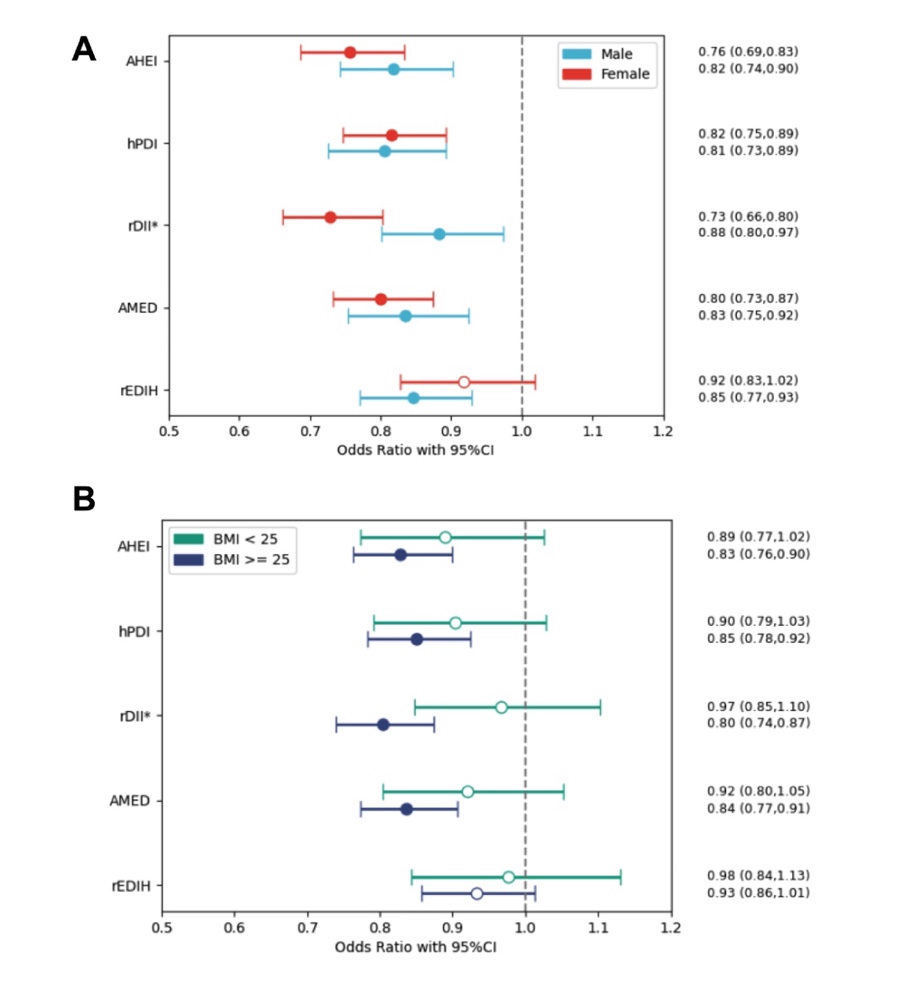

### Supplementary Figure 5. Association of adherence to different healthy dietary patterns with prevalent MASLD risk.

The odds ratios (OR, solid line) and 95% confidence intervals (CIs, shade area) were estimated with logistic regression model with a restricted cubic spline (RCS) using three knots (10^th^, 50^th^ and 90^th^ percentiles of dietary scores). The minimum value of each dietary pattern score was used as a reference value. AHEI, Alternative Healthy Eating Index; AMED, Alternate Mediterranean Diet Score; hPDI, healthy Plant-based Diet Index; rDII, reversed Dietary Inflammatory Index; rEDIH, reversed Empirical Dietary Index for Hyperinsulinemia.

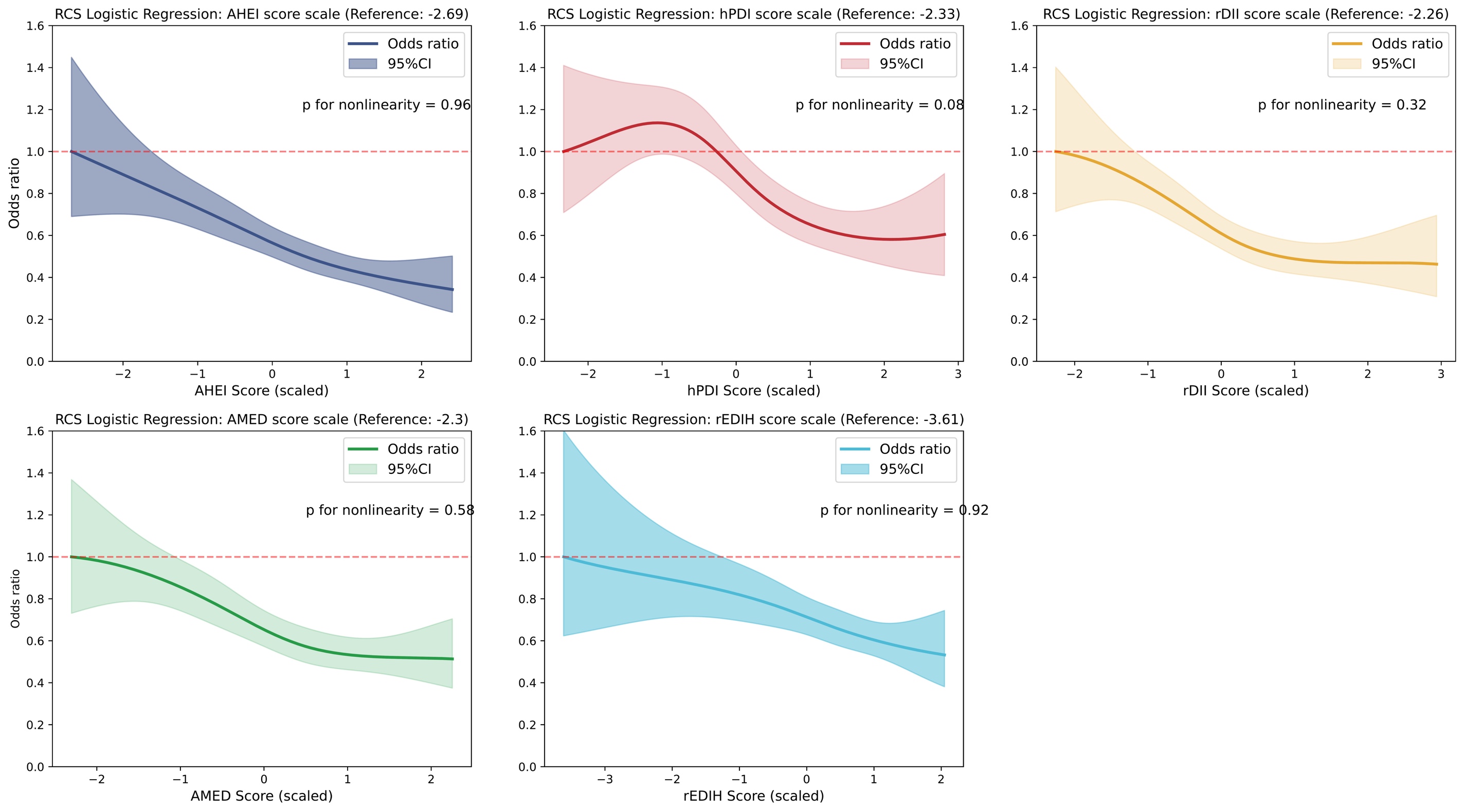

### Supplementary Figure 6. Association of each dietary patterns score (quintiles) and prevalent MASLD risk.

The odds ratios (ORs) and 95% confidence intervals (CIs) were estimated using logistic regression models adjusted for age, sex, education level, smoking status, sleep duration, physical activity, vitamin use, and hormone use. An additional adjustment for alcohol intake was made when hPDI served as the exposure. The lowest quintile (Q1) of each dietary pattern score served as the reference group. Solid dots indicate statistically significant associations (*P* < 0.05); hollow dots indicate non-significant associations. AHEI, Alternative Healthy Eating Index; AMED, Alternate Mediterranean Diet score; hPDI, healthy Plant-based Diet Index; rDII, reversed Dietary Inflammatory Index; rEDIH, reversed Empirical Dietary Index for Hyperinsulinemia.

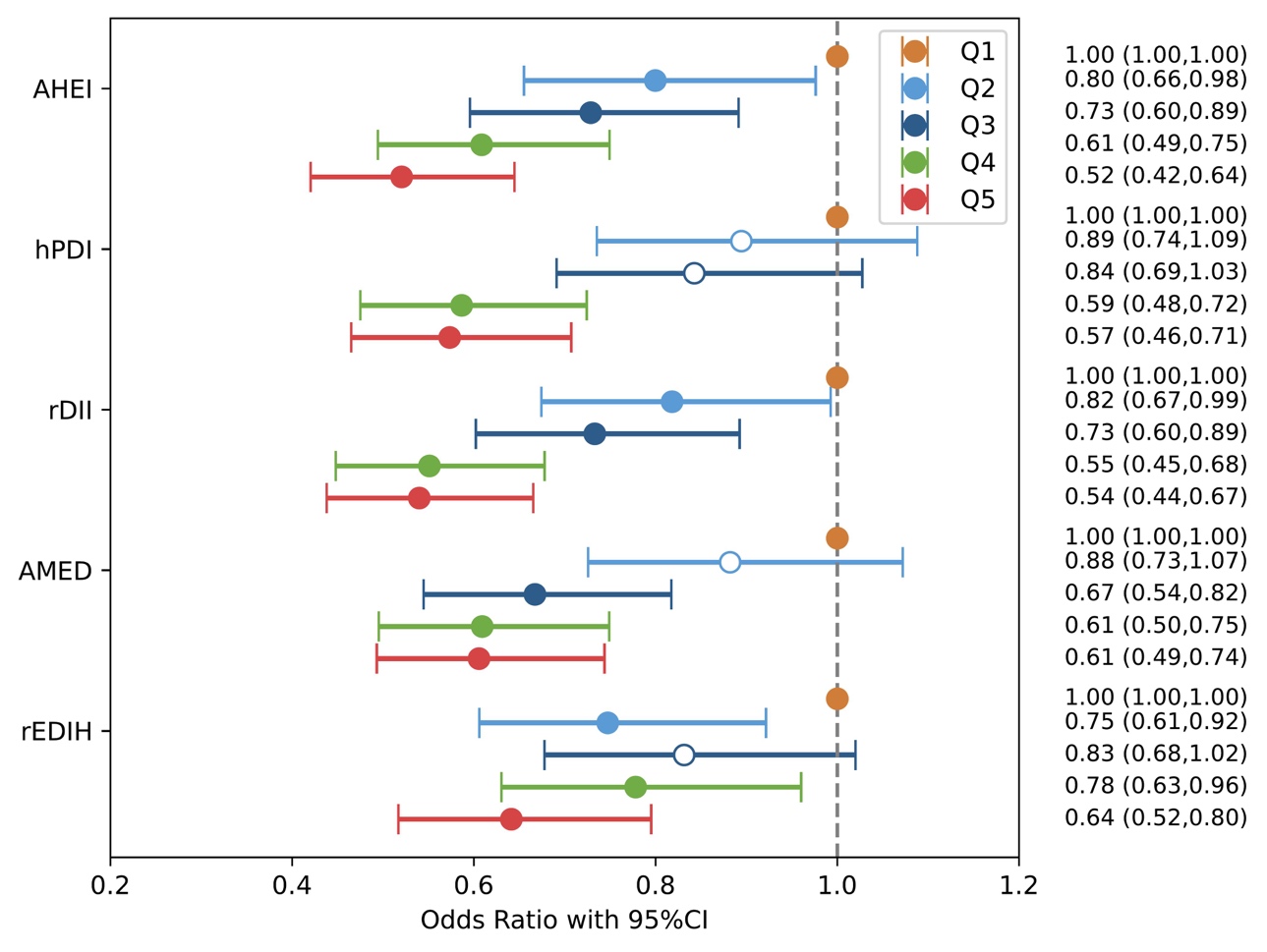

### Supplementary Figure 7. Sensitivity analyses for the associations between adherence to different dietary patterns and self-reported prevalent MASLD.

The odds ratios (ORs) and 95% confidence intervals (CIs) were estimated using logistic regression models. Four sequential models were fitted: Model 1, unadjusted; Model 2, adjusted for age, sex, education level, smoking status, sleep duration, physical activity, vitamin use, and hormone use; Model 3, Model 2 plus BMI; Model 4, Model 2 plus NSAID use, family history of diabetes, and family history of CVD. An additional adjustment for alcohol intake was made when hPDI served as the exposure. Solid dots indicate statistically significant associations (*P* < 0.05); hollow dots indicate non-significant associations. AHEI, Alternative Healthy Eating Index; AMED, Alternate Mediterranean Diet score; BMI, body mass index; CVD, cardiovascular disease; hPDI, healthy Plant-based Diet Index; MASLD, metabolic dysfunction-associated steatotic liver disease; NSAID, nonsteroidal anti-inflammatory drugs; rDII, reversed Dietary Inflammatory Index; rEDIH, reversed Empirical Dietary Index for Hyperinsulinemia.

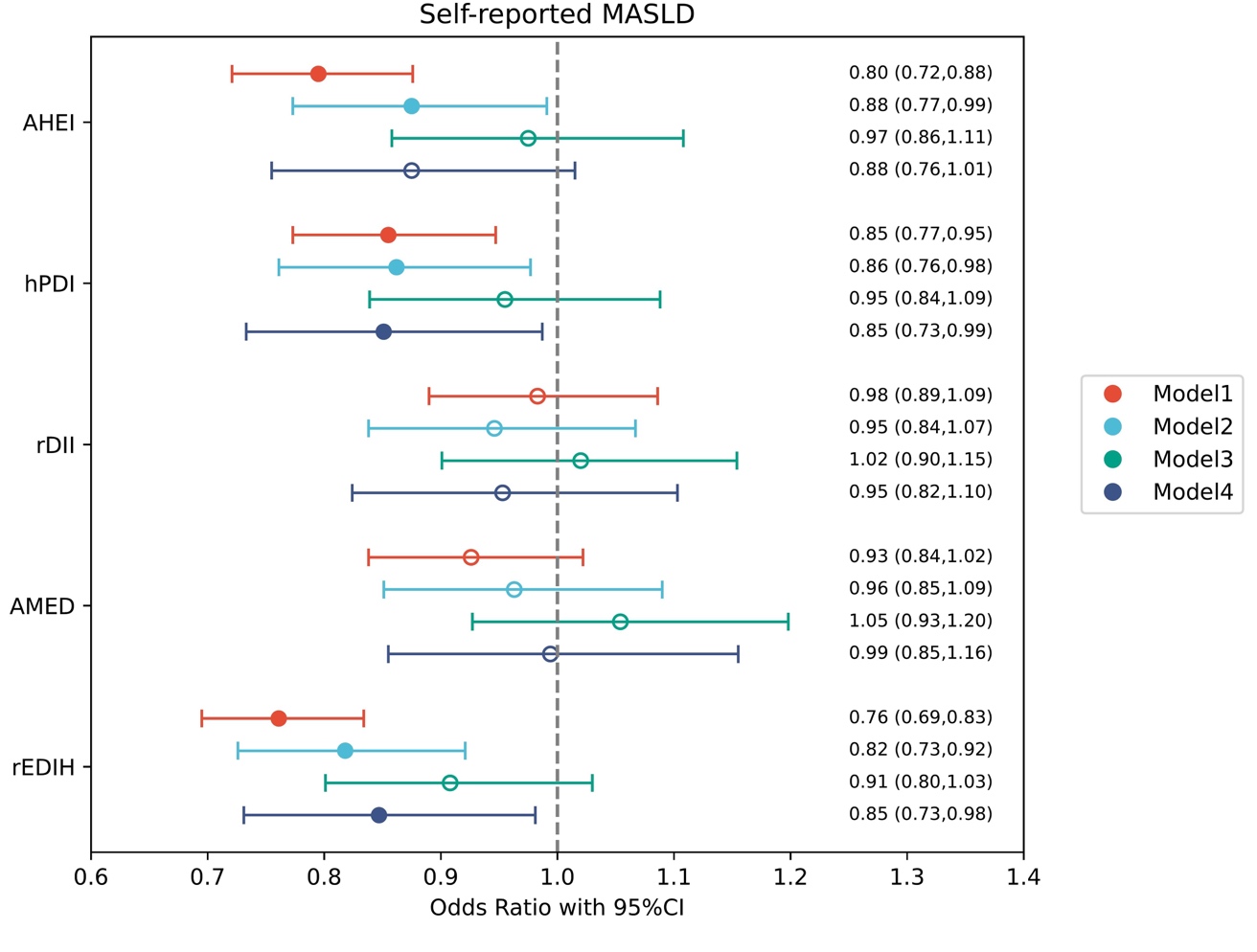

### Supplementary Figure 8. Associations between adherence to healthy dietary patterns and hepatic metrics from liver ultrasound and 2D-SWE, stratified by sex and BMI.

Forest plots show β coefficients with 95% confidence intervals (CIs) per 1-SD increase in dietary pattern scores for associations with six hepatic metrics (speed of sound, attenuation coefficient, liver elasticity, velocity, viscosity, and dispersion), stratified by sex (**A**) and BMI category (<25 vs. ≥ 25 kg/m²; **B**). β coefficients were obtained from generalized linear models. In **A**, models were adjusted for age, education level, smoking status, sleep duration, physical activity, vitamin use, and hormone use. In **B**, models were adjusted for the same covariates plus sex. For hPDI, models were additionally adjusted for alcohol intake. The solid dots indicate statistically significant associations (*P* < 0.05); the hollow points indicate non-significant association. AHEI, Alternative Healthy Eating Index; AMED, Alternate Mediterranean Diet Score; BMI, body mass index; FDR, false discovery rate; hPDI, healthy Plant-based Diet Index; rDII, reversed Dietary Inflammatory Index; rEDIH, reversed Empirical Dietary Index for Hyperinsulinemia; 2D-SWE, two-dimensional shear wave elastography.

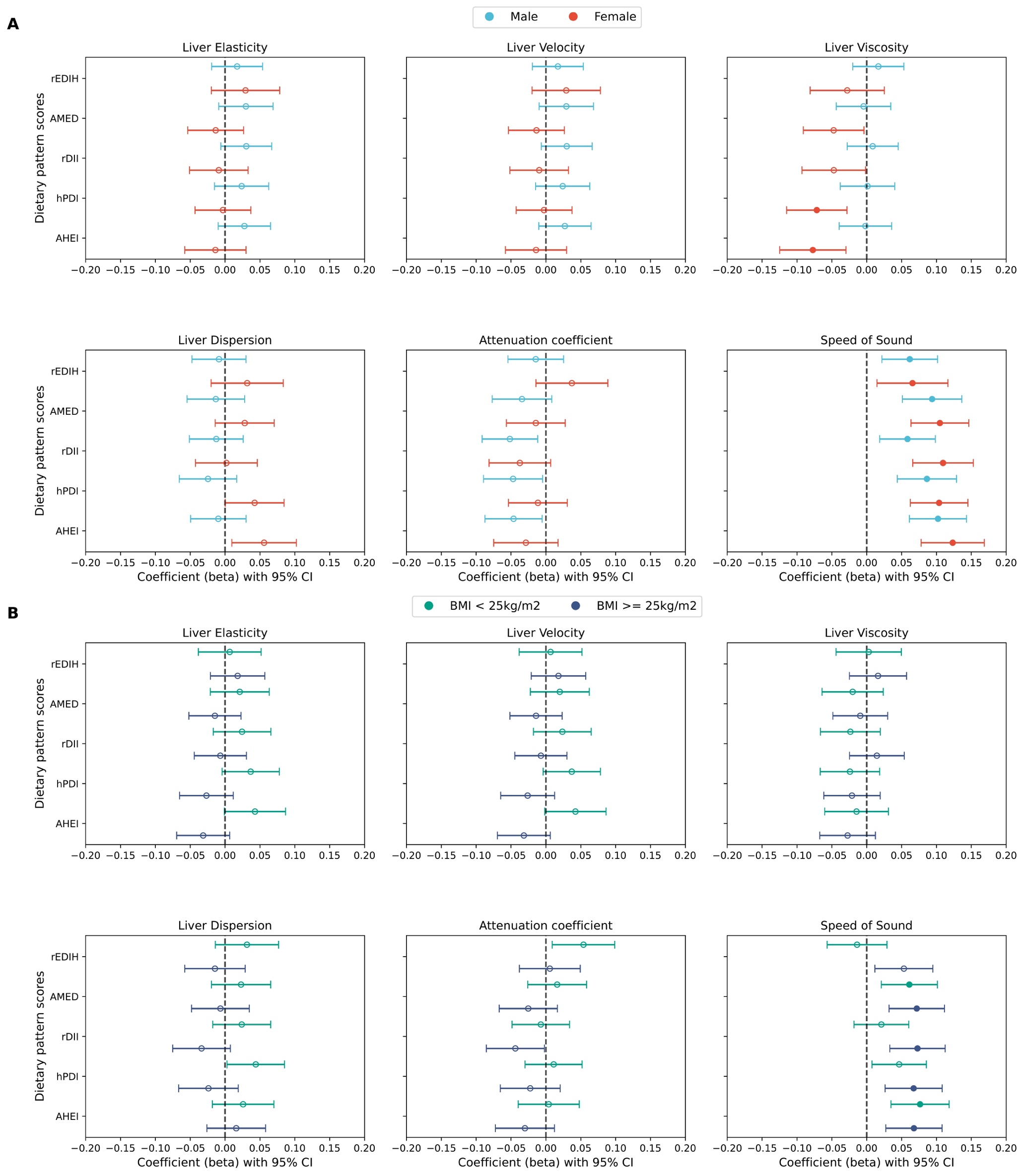

### Supplementary Figure 9. Associations between adherence to different dietary patterns (quintiles) and hepatic metric obtained from 2D-SWE.

The β coefficients with 95% confidence intervals (CIs) were estimated using linear regression models adjusted for age, sex, education level, smoking status, sleep duration, physical activity, vitamin use, and hormone use. An additional adjustment for alcohol intake was made when hPDI served as the exposure. The lowest quintile of each dietary pattern score served as the reference group. AHEI, Alternative Healthy Eating Index; AMED, Alternate Mediterranean Diet score; hPDI, healthy Plant-based Diet Index; rDII, reversed Dietary Inflammatory Index; rEDIH, reversed Empirical Dietary Index for Hyperinsulinemia.

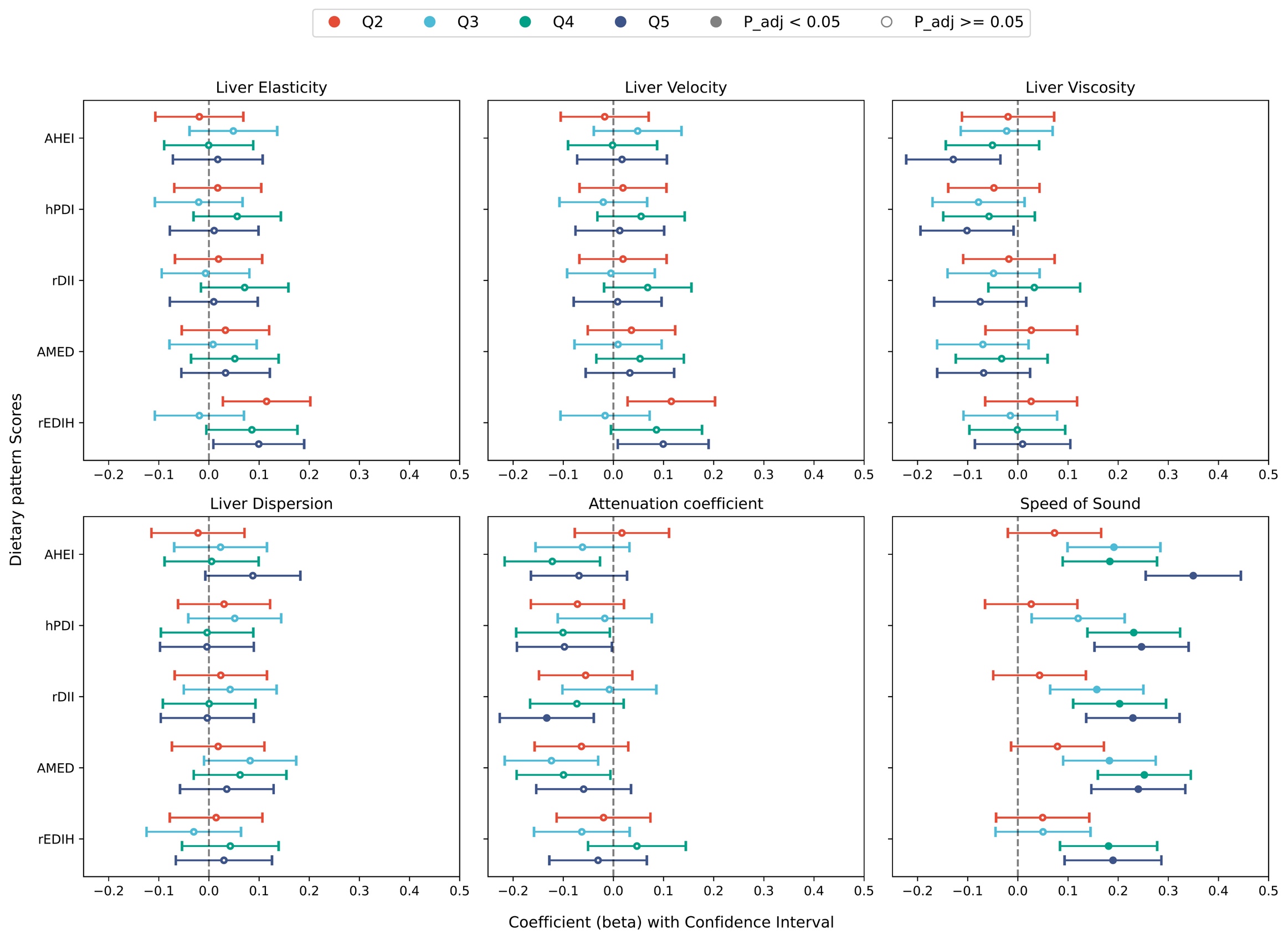

### Supplementary Figure 10. Sensitivity analyses for the associations between adherence to different dietary patterns and hepatic metrics derived from liver ultrasound and 2D-SWE.

The β coefficients with 95% confidence intervals (CIs) were estimated using linear regression models. (**A**) Results after truncating hepatic metrics at the 1st and 99th percentiles to minimize the influence of measurement outliers. (**B**) Results after excluding individuals whose liver speed of sound (SoS) exceeded the 99th percentile. Four sequential models were fitted: Model 1, unadjusted; Model 2, adjusted for age, sex, education level, smoking status, sleep duration, physical activity, vitamin use, and hormone use; Model 3, Model 2 plus BMI; Model 4, Model 2 plus NSAID use, family history of diabetes, and family history of CVD. An additional adjustment for alcohol intake was made when hPDI served as the exposure. Solid dots represent associations that remained significant after correction for multiple comparisons (*FDR*-adjusted *P* < 0.05). AHEI, Alternative Healthy Eating Index; AMED, Alternate Mediterranean Diet score; BMI, body mass index; CVD, cardiovascular disease; hPDI, healthy Plant-based Diet Index; NSAID, nonsteroidal anti-inflammatory drugs; rDII, reversed Dietary Inflammatory Index; rEDIH, reversed Empirical Dietary Index for Hyperinsulinemia.

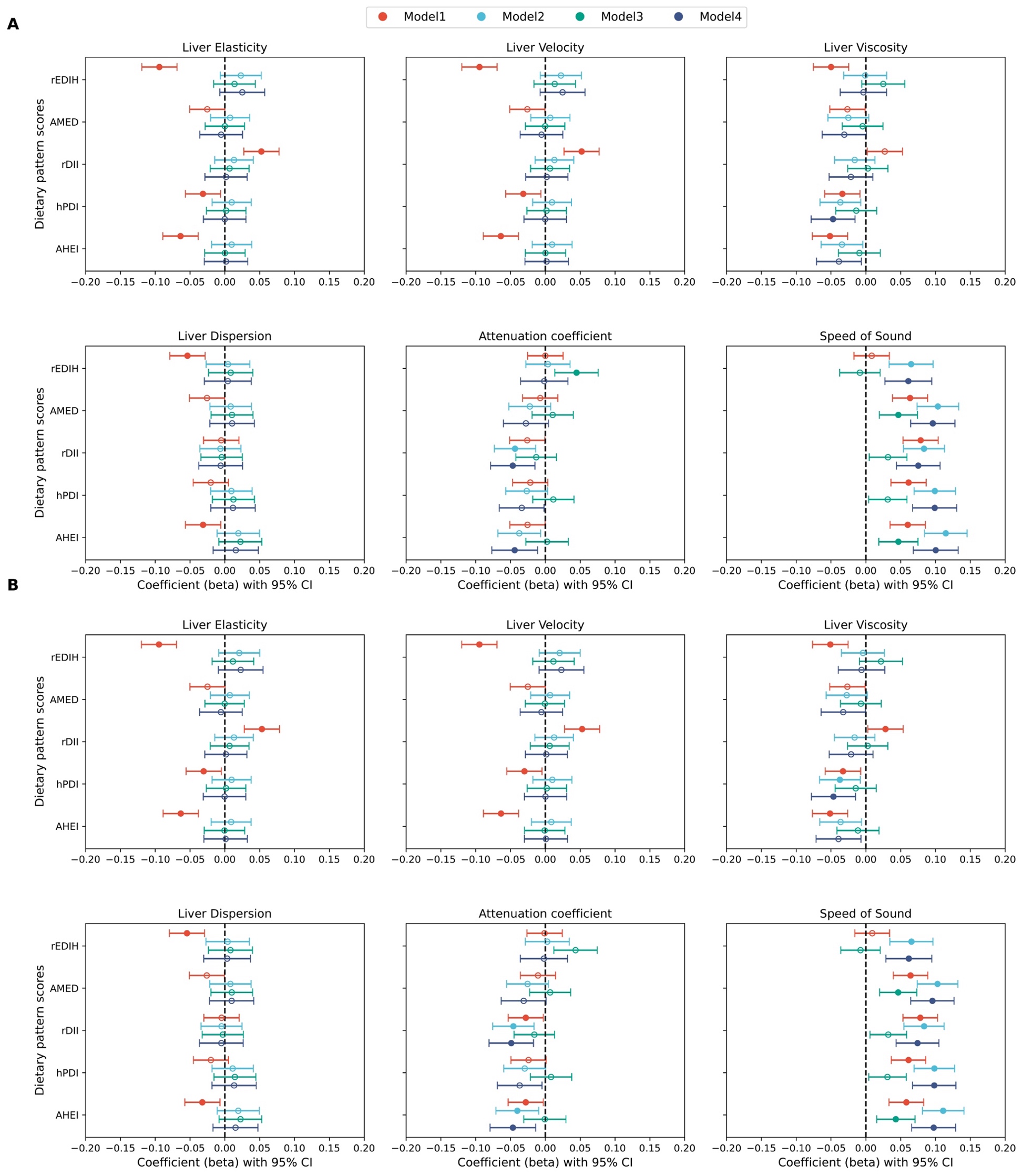

### Supplementary Figure 11. Associations between healthy dietary pattern adherence and overall gut microbial composition.

(**A**) Composition of the most abundant phyla in the study population. (**B**) Shannon index and Simpson index across quintiles of five dietary pattern scores (Kruskal–Wallis test). (**C**) Principal coordinates analysis (PCoA) based on species-level Bray–Curtis dissimilarity, showing the association between microbial composition and adherence to different healthy dietary patterns (PERMANOVA, 999 permutations). AHEI, Alternative Healthy Eating Index; AMED, Alternate Mediterranean Diet score; hPDI, healthy Plant-based Diet Index; rDII, reversed Dietary Inflammatory Index; rEDIH, reversed Empirical Dietary Index for Hyperinsulinemia.

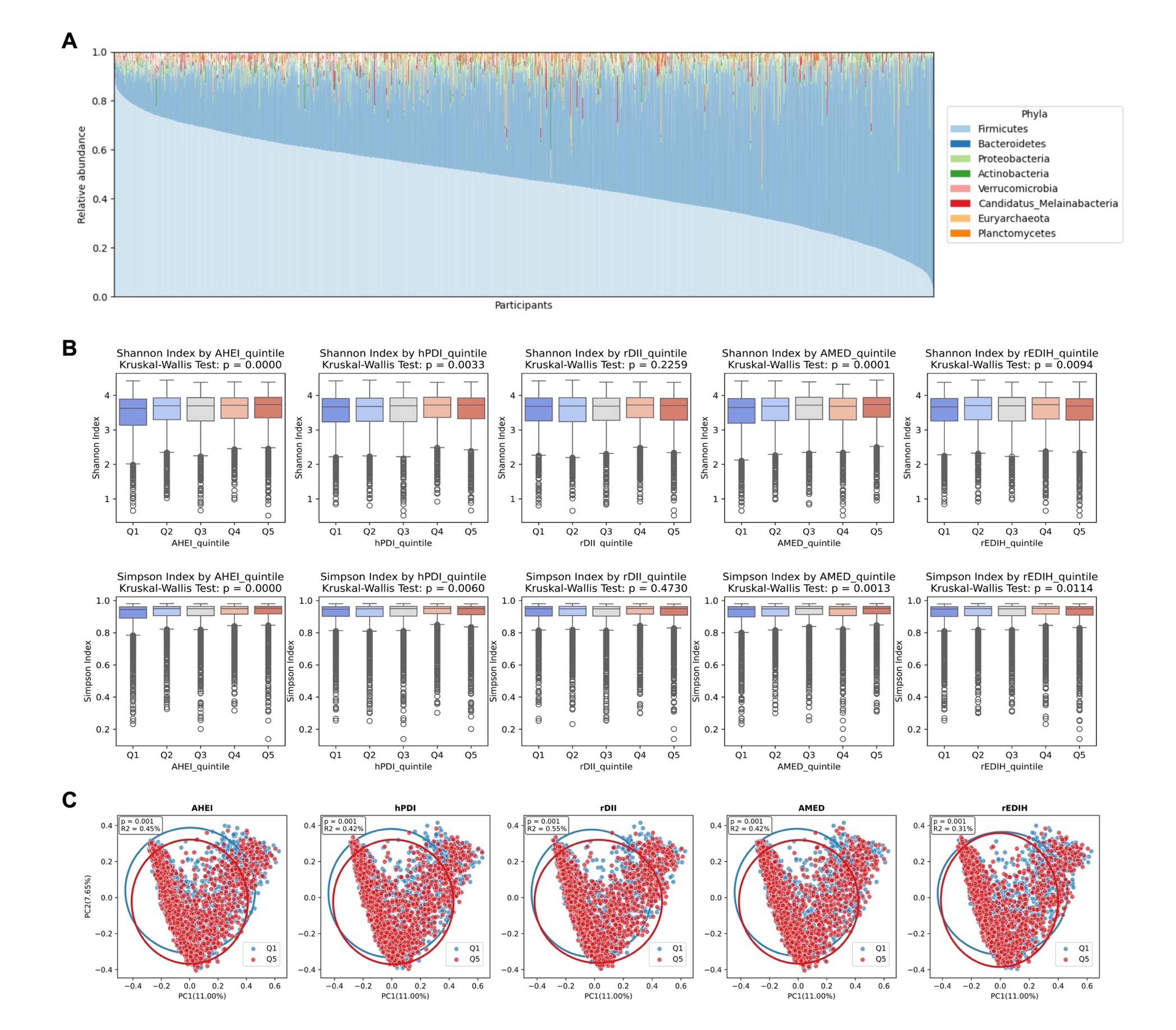

### Supplementary Figure 12. UpSet plot showing the numbers of shared and exclusive healthy diet-associated microbial species across different dietary pattern scores.

In the intersection matrix, filled (colored) dots indicate dietary patterns included in each intersection, connected by lines for multi-pattern intersection, whereas grey dots indicate dietary patterns not included. The horizontal bars on the right show the total number of gut microbial species associated with each dietary pattern score. In the primary analysis, out of 379 microbial species, there were 185, 250, 251, 256, and 261 species associated with adherence to rEDIH, AHEI, AMED, rDII, and hPDI in the same direction of effect, respectively: 138 microbial species showed significant association with adherence to all five dietary patterns, while there were 2, 5, 6, and 17 microbial species showing exclusive associations with AHEI, hPDI, rDII, and rEDIH. AHEI, Alternative Healthy Eating Index; AMED, Alternate Mediterranean Diet Score; hPDI, healthy Plant-based Diet Index; rDII, reversed Dietary Inflammatory Index; rEDIH, reversed Empirical Dietary Index for Hyperinsulinemia.

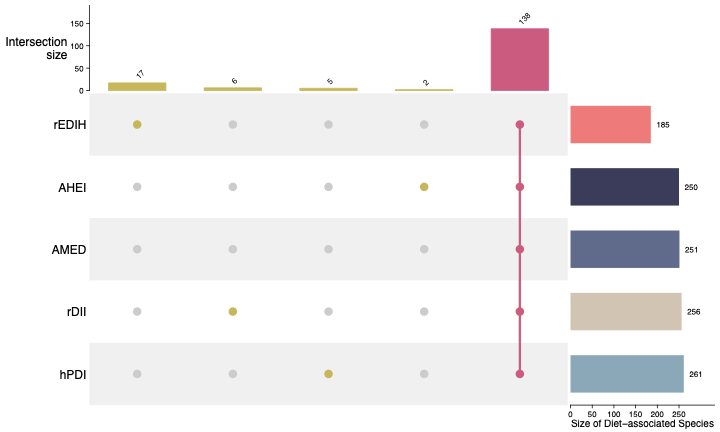

### Supplementary Figure 13. Associations between exclusive diet-associated microbial species and hepatic metrics derived from liver ultrasound and 2D-SWE.

The heatmap shows associations between microbial species that were exclusively associated with a single healthy dietary pattern score and hepatic metrics derived from liver ultrasound and 2D-SWE. The values in the cells indicate the β coefficients obtained from linear regression models, with each hepatic metric as the dependent variable and each microbial species as the independent variable, adjusted for age, sex, BMI, education level, smoking status, sleep duration, physical activity, alcohol intake, vitamin use, hormone use, and antibiotic and proton pump inhibitor (PPI) use during the dietary logging period. Values marked with an asterisk denote associations that remained statistically significant after correction for multiple testing using the Benjamini–Hochberg method (*FDR*-adjusted *P* < 0.05).

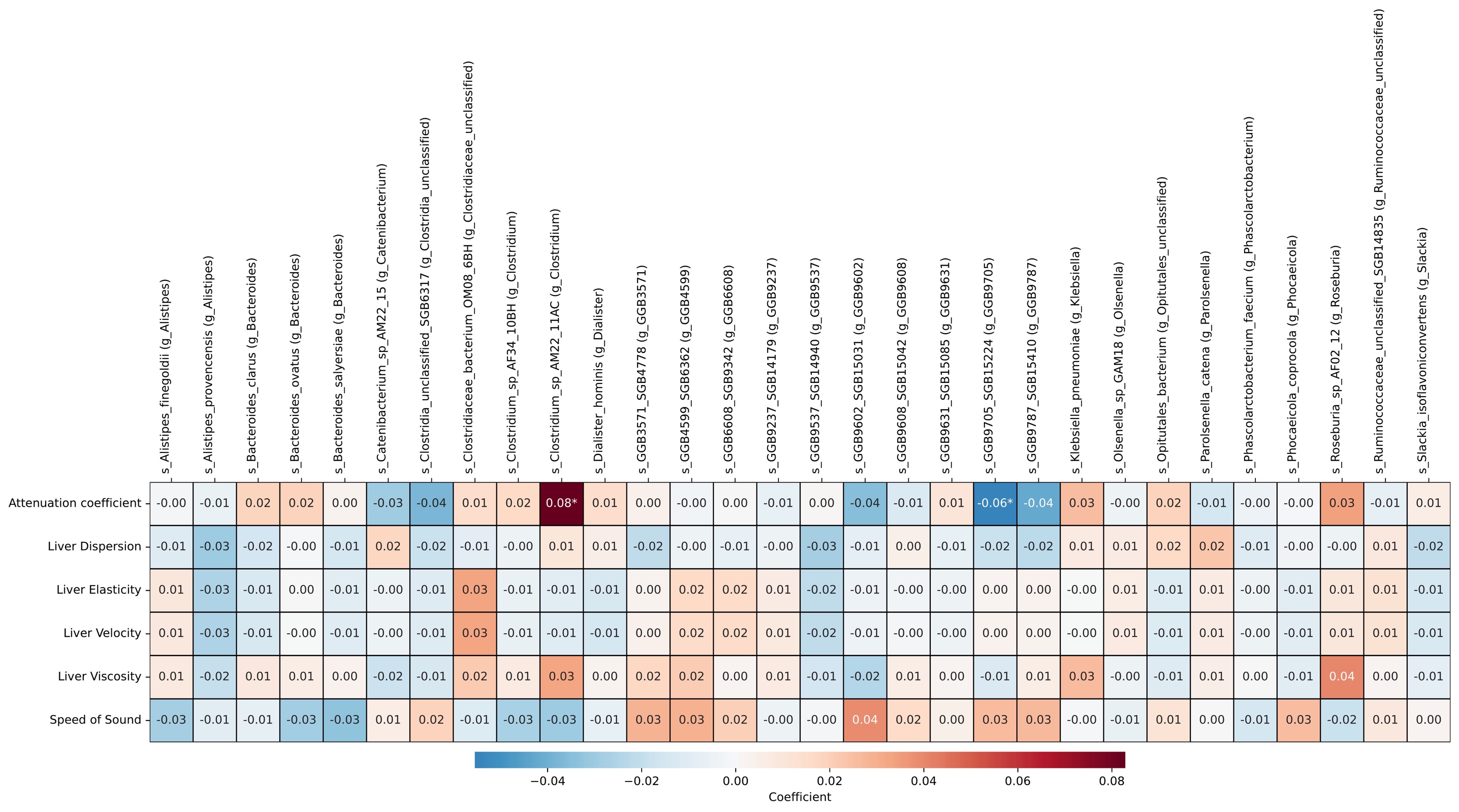

### Supplementary Figure 14. Associations between exclusive diet-associated microbial species and prevalent MASLD risk.

The forest plot shows adjusted odds ratios (ORs) and 95% confidence intervals (CIs) for associations between microbial species that were exclusively associated with a single healthy dietary pattern score and prevalent MASLD. The ORs and 95% CIs were obtained from logistic regression models with MASLD prevalence as the dependent variable and each microbial species as the independent variable, adjusted for age, sex, BMI, education level, smoking status, sleep duration, physical activity, alcohol intake, vitamin use, hormone use, and antibiotic and proton pump inhibitor (PPI) use during the dietary logging period. Grey points indicate associations that were not statistically significant after correction for multiple testing using the Benjamini–Hochberg method (*FDR*-adjusted *P* > 0.05).

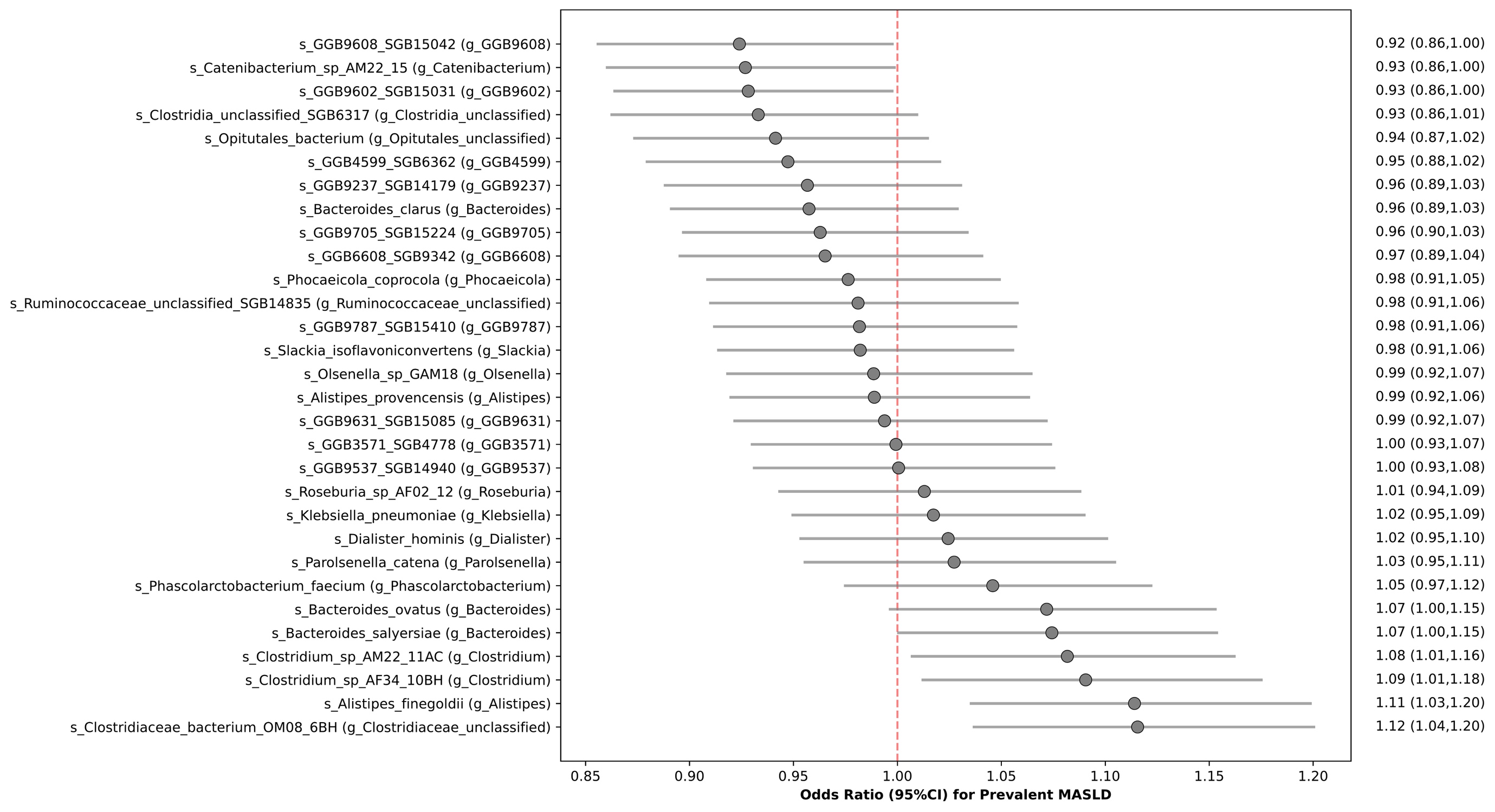

### Supplementary Figure 15. Mediating roles of microbial species in the association between healthy dietary pattern adherence and prevalent MASLD.

Cross-sectional mediation analyses were conducted to examine whether microbial species statistically mediate the association between healthy diet adherence and prevalent MASLD. The forest plots display the estimated proportion mediated by 20 microbial species in the association between greater adherence to healthy dietary patterns and prevalent MASLD. The analyses were adjusted for age, sex, BMI, education level, smoking status, sleep duration, physical activity, vitamin use, hormone use, and antibiotic and proton pump inhibitor (PPI) use during the dietary logging period. An additional adjustment for alcohol intake was made when hPDI served as the exposure. The 95% confidence intervals (CIs) of the mediated proportion were derived using 1,000 bootstrap iterations.

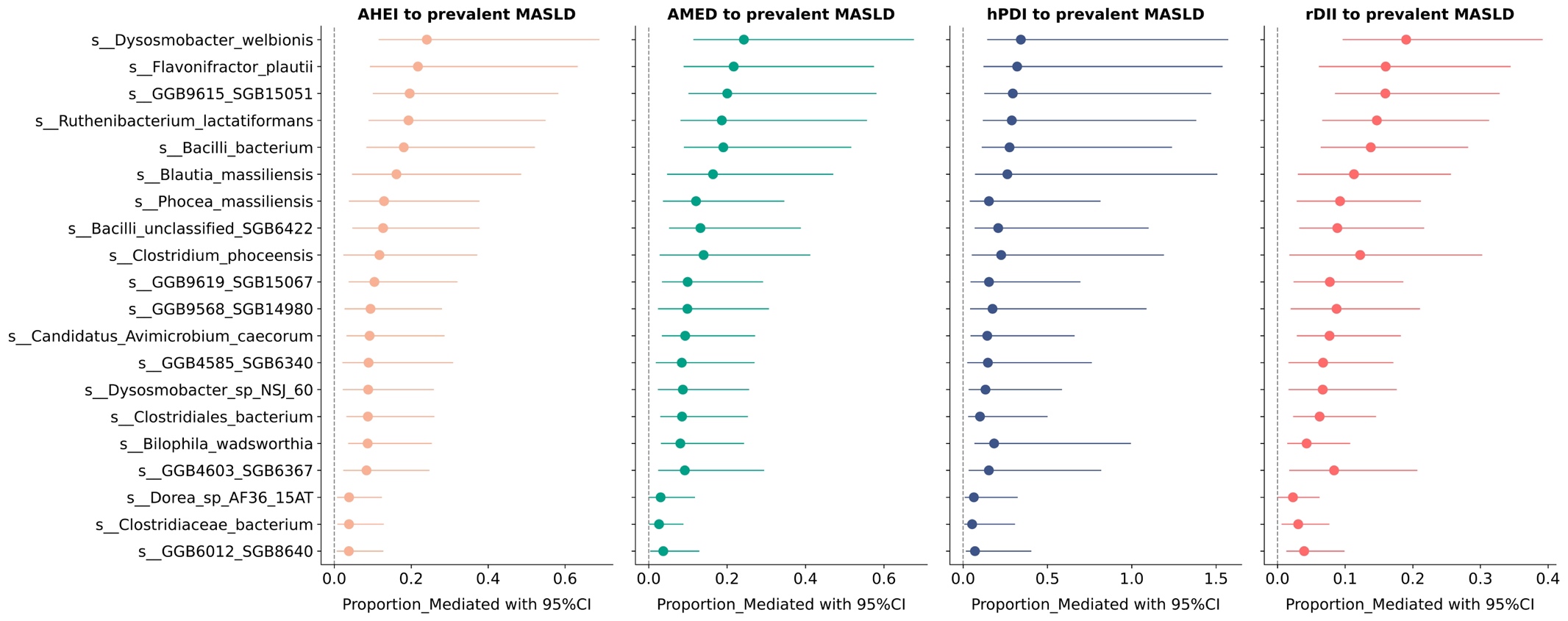

### SUPPLEMENTARY TABLES

**Supplementary Table 1.** Associations between adherence to healthy dietary patterns and hepatic metrics from ultrasound.

**Supplementary Table 2.** The explained variance of Shannon index by different dietary pattern scores.

**Supplementary Table 3.** Associations between adherence to healthy dietary patterns and abundance of gut microbial species (Model 2).

**Supplementary Table 4.** The number of exclusive associations of adherence to healthy dietary patterns with abundance of gut microbial species.

**Supplementary Table 5.** Associations between adherence to healthy dietary patterns and abundance of gut microbial species (Model 3 and 4).

**Supplementary Table 6.** Consistent associations between adherence to healthy dietary patterns and abundance of gut microbial species across all tested models.

**Supplementary Table 7.** Associations between diet-related gut microbial species and hepatic outcomes.

**Supplementary Table 8.** Mediating roles of microbial species between healthy diet adherence and liver speed of sound.

**Supplementary Table 9.** Components of the Alternate Mediterranean Diet (AMED) scoring method.

**Supplementary Table 10.** Components of the Alternate Healty Eating Index (AHEI) scoring method.

**Supplementary Table 11.** Examples of food items constituting the 18 food groups and healthy Plant-based Dietary Index (hPDI) scoring method.

**Supplementary Table 12.** Components of the Empirical Diet Index for Hyperinsulinemia (EDIH) scoring method.

**Supplementary Table 13.** Food components included in the Dietary Inflammatory Index (DII), inflammatory effect scores, and intake values from the global composite date set.
